# Impacts of British Columbia’s free contraception policy on out-of-pocket payments and contraception costs: an interrupted time series analysis with other provinces as controls

**DOI:** 10.1101/2025.11.04.25339524

**Authors:** Elizabeth Nethery, Michael R Law, Lucy Chang, Stirling Bryan, Fiona Clement, Nathan Nickel, Wendy V Norman, Amanda Black, Mary Helmer-Smith, Kimberlyn M McGrail, Laura Schummers

## Abstract

**Background:** Cost is a key driver of inadequate, unequal contraception access. Without a national public pharmacare plan, Canadians use a mix of private or public prescription insurance plans to pay for drugs or devices. High deductibles, copayments, and/or exclusion of contraception from insurance formularies can result in access gaps. In April 2023, the western-most Canadian province (British Columbia (BC)) introduced universal, first-dollar coverage for prescription contraception. We estimated the effect of this free contraception policy on prescription contraception payer type and costs.

**Methods:** Using a national prescription database, we measured the monthly proportion of all dispensed contraception to reproductive-aged females (15-49y) in BC according to majority payer (out-of-pocket (patients), private, and public) and estimated costs per capita by payer type. We used a controlled interrupted time series analysis to compare outcomes in BC to expected trends if the policy had not been introduced and compared with synthetic controls derived from other Canadian provinces.

**Findings:** Pre-policy in BC, 39% of prescription contraception was paid out-of-pocket, 49% by private insurance, and 12% by public insurance. When BC’s policy was introduced, out-of-pocket payments immediately decreased (−24.3% 95%CI −24.8 to −23.7). At 2-years post-policy, 5.4% (95% CI 4.5% to 6.4%) of on-formulary contraceptives were paid out-of-pocket and 8.4% (7.5% to 9.3%) was paid by private insurance. Total monthly per capita out-of-pocket costs in BC were 86% lower than expected if the policy had not been implemented. Time trends pre-and post-policy were similar in BC and controls.

**Interpretation:** Up to 40% of contraceptives were previously paid out-of-pocket in BC compared to prior studies showing 20% of all drugs in Canada were paid out-of-pocket. Contraception may be uniquely sensitive to inadequate insurance coverage as with a mix of public and private drug plans. The implementation of a public drug plan providing universal, first-dollar coverage policy for contraception substantially reduced out-of-pocket payments for dispensed contraception.

**Key points:**

- Before policy implementation, 40% of contraception prescriptions in BC were paid out-of-pocket – double the national average for all prescription drugs – highlighting major gaps in coverage and inequitable access to contraception.
- A universal, first-dollar contraception policy reduced out-of-pocket costs by 86% within two years, with only 5% of on-formulary contraceptive prescriptions still paid out-of-pocket
- First-dollar public coverage effectively removes financial barriers and supports equitable access to contraception, providing timely evidence to inform national pharmacare implementation.

## BACKGROUND

Access to effective contraception is essential to enable pregnancy-capable individuals to control timing and spacing of pregnancies.^1^ Approximately 40% of pregnancies in Canada are unintended, and, while not all are undesired, these are associated with increased costs to the health system.^2^ Optimal pregnancy timing improves health outcomes,^3^ reduces health inequities and poverty ^4–6^, and improves educational attainment and economic gender equity.^6–8^ However, high costs of contraception remains a key driver of inadequate, unequal contraception access in Canada.^9–12^ In 2016, the UN Human Rights Commissioner called on Canada to improve access to affordable contraception.^4^ Prescription contraception costs range from an up-front payment of $350-$400 for the most effective contraception methods (long-acting reversable contraceptives (LARC) such as hormonal intrauterine devices (IUDs) (lasting 5 to 8 years) or subdermal contraceptive implants (3 years)) to $30 per month for short-acting reversable contraceptives (SARC) (e.g. oral contraceptive pills, contraceptive ring, patch or injectables). Cost poses a significant access barrier.^9,12,13^ High prescription contraception costs and poor contraceptive adherence are among the reasons that unintended pregnancy is more common in lower income groups.^2,14–16^

Canada remains one of the only high-income nations with single-payer (provincial) health systems that did not universally cover contraception. While a national Pharmacare Act^17^ was passed on October 10, 2024, only 3 provinces and 1 territory have agreements with the federal government and/or have implemented universal contraceptive coverage. Before the national act was passed, the province of British Columbia (BC)^18,19^ implemented Canada’s first universal, first-dollar contraception subsidy on April 1, 2023, making most prescription contraception (and all LARC) free for provincially insured residents. The policy aim was improved access to contraception to achieve reproductive health and equity.^18,20^ Following BC’s lead, Manitoba announced a similar contraception subsidy that took effect on October 1, 2024. Bilateral federal-provincial/territorial agreements to implement the Pharmacare Act^21,22^ have been signed in 2 other regions (the Yukon and Prince Edward Island). While most Canadians have some form of private or public insurance for medication, many face significant out-of-pocket costs due to high deductibles and/or copayments.^23,24^ In addition to cost barriers, concerns about health privacy, specific to contraception, may create access barriers because private insurance plans are often shared with family members.^25,26^ A universal, first-dollar insurance plan means that all eligible people are covered from the first dollar of cost (i.e. no deductibles or co-payments). Furthermore, because BC’s public insurance drug coverage is implemented via the public medical system – prescription records subject to health privacy laws and may only be accessed by the patient or authorized health care professionals. As the first Canadian province to implement contraceptive coverage through a universal plan, BC provides a unique opportunity to characterize changes for the provincial (public) insurance payer, contraception users paying out-of-pocket, and private insurance plans. Therefore, we estimated the effect of this policy on payer type and direct costs for dispensed prescription contraception.

## METHODS

### Setting

Nearly all Canadian residents have health insurance coverage through single-payer provincial insurance plans (or federal programs, e.g., military/federal police, legal refugee/asylum seekers, some First Nations people); however, prescription drug coverage is not universally included.^23,27^ Most provinces have a public drug plan which also requires patients to pay out-of-pocket through copayments or income-based deductibles.^28^ As of 2024, public plans were estimated to cover 42% of prescription costs in Canada, private plans (mainly funded by employers) were estimated to cover 38%, and the remaining 20% was paid out-of-pocket by patients.^29,30^

In BC, the public health insurance plan (BC Medical Services Plan (MSP)) covers all eligible BC residents and dependents (Canadian citizens or permanent residents who reside in BC at least 6 months per year) as well as some individuals on study or work permits. Coverage begins following a 3-month waiting period from the date of application. BC’s public drug plan (BC PharmaCare) includes 12 sub-plans with specific eligibility criteria (e.g., income assistance, long-term care residents). One of these plans (Assurance/Plan Z) automatically provides 100% coverage to all BC residents with MSP for specific drugs: including mifepristone for medication abortion, opioid use disorder treatment, and medications used for medical assistance in dying.^31^ Manitoba has a similar public insurance plan (Manitoba Health).

### Intervention

On April 1, 2023, BC introduced universal prescription contraception by adding specified contraceptives to the existing Plan Z formulary.^32^ This provided 100% coverage for all LARC (copper and levonorgestrel-releasing IUDs, levonorgestrel-releasing contraceptive implants) and injectable contraception, and partial coverage up to generic price for brand-names for the contraceptive vaginal ring, most oral contraceptive pill (OCP) formulations, and emergency contraceptives. BC also implemented Exceptional Plan Z coverage for those in the waiting period for MSP enrollment^31^ and two OCPs were later added to the formulary (Supplemental Table 1). The province of Manitoba also implemented a program to cover 100% of the costs for prescription contraceptives (Manitoba Prescription Birth Control Program) on October 1, 2024 using the same formulary as BC for anyone with a Manitoba Health Card (ie. active provincial insurance).

### Study design

We used a controlled interrupted time series (CITS) study design to evaluate the impact of contraception coverage on payer type and costs for prescription contraception in BC (the intervention group), using a synthetic control ^33,34^ derived from Canadian provinces without universal contraception coverage, from April 1, 2021 to March 31, 2025. Interrupted time series (ITS) is a quasi-experimental design that analyzes expected outcome frequencies before and after an intervention or policy change ^35,36^. This design extends pre-intervention trends to derive a counterfactual in the post-intervention period and uses statistical models estimate a level change (an immediate increase or decrease) and a trend change at the time point of the intervention. ITS analyses are generally not vulnerable to confounding by individual-level covariates, but can be impacted by other events that co-occur with the policy change ^35^. By using a control group which is not exposed to the policy of interest, we improve the validity by controlling for any possible co-interventions (other events or policy changes) that may impact study outcomes ^33,37^. In a CITS design, the counterfactual is also informed by changes in the outcome level or trend occurring in the control group. As a secondary analysis, we also evaluated the impact of a free contraception policy in Manitoba using the same approach.

### Data source

Our study used the IQVIA Geographic Prescription Monitor database that contains outpatient pharmacy transactions for all dispensed pharmaceutical products across Canadian provinces and in subgroups by age and primary payer type (cash, public payer, private insurance). Using a panel of retail pharmacies capturing approximately 83% of all prescriptions dispensed (per 2024 sample coverage) as the sample frame, this database estimates all (100%) dispensations at Canadian pharmacies using geospatial projections with established validity ^38^ and prior use in research ^39–41^. We previously compared dispensation volumes in this database with BC population-based administrative health data and found nearly identical volumes of contraception prescriptions and matching trends throughout our study period ^42^. We examined contraception dispensations to reproductive-aged (15-49y) females (administrative gender ^43^) in the 10 Canadian provinces.

### Outcomes

We examined two primary outcomes: 1) the proportion of contraception dispensations by payer group (out-of-pocket [patients], public and private insurance) and 2) the costs of dispensed contraception per capita. Costs were based on the number of units dispensed multiplied by current drug unit costs, an 8% markup and addition of a per-dispensation cost ($10) per BC policies ^44^. Costs ($ CAD) per capita used monthly provincial populations of reproductive aged females (15 to 49y) and within appropriate age-strata ^45^.

### Statistical analysis

The impact of BC’s free contraception policy (April 1, 2023) was modeled using CITS analysis comparing monthly rates for payer type proportions and costs per capita (out-of-pocket, public, and private insurance). We also examined out-of-pocket costs per capita in age subgroups (15-19y, 20-29y, 30-39y, 40-49y), total costs per capita (all payers) for all contraceptives and by contraception type (SARC and LARC) and total costs by payer. We used the R(Synth) package to derive weights for each potential control province (other provinces without universal contraception coverage) ^46^.

Generally, this approach constructs a single synthetic control using a generalization method for a typical differences-in-differences model that identifies an optimal set of weights to minimize the mean squared error of the pre-policy difference between the synthetic control and intervention group ^34^. The resulting weights (from 0 to 1 for each province) are chosen for each CITS model so the resulting synthetic control best represents the pre-policy level and trends. Models included parameters for group (BC vs. synthetic control) and interaction terms for pre-policy trend and group (pre-policy slope in BC vs. control), level and group (level change in BC vs. control at the intervention time), and post-policy trend and group (post-policy trend change in BC vs. control). We modeled the month of April 2023 as a phase-in period as this first post-policy month showed a spike in most outcomes, indicating pent-up demand and change in contraception use patterns in anticipation of the policy introduction ^47^. Our models used a Prais-Winsten method which produces a single measure of autocorrelation, is unbiased compared to an ARIMA model, and is recommended for analyzing ITS ^48,49^. We added model parameters for appropriate months (seasonality) for any observed spikes, following best practices in ITS modeling ^50^. We used the resulting model estimates to estimate absolute (predicted level minus counterfactual level) and relative (predicted level divided by counterfactual level) policy effects at 2-years post-policy (March 2025). Bootstrapping with 1,000 iterations estimated confidence intervals around these summary measures ^51^. Models for Manitoba’s policy (October 1, 2024) excluded BC as a potential control province and BC models excluded Manitoba. All analyses were conducted in SAS version 9.4 and R Version 4.4.3. ^52,53^ We received Research Ethics Board approval from the University of British Columbia (H23-04138).

## RESULTS

From April 2021 to the implementation of free contraception in BC in April 2023 (24 months), contraceptive dispensations by payer type were generally stable in all 10 provinces (Figure 1, A-C). In the pre-policy period, BC had the highest proportion of out-of-pocket payments (39% of monthly dispensations for prescription contraception), while 49% had private insurance, and 12% were paid by public insurance. Immediately following the policy change in BC, there was an estimated immediate decrease in the proportion of dispensations paid out-of-pocket (level change: −24.3%, 95% CI −24.8 to −23.7) and by private insurance (level change: −35.9%, 95% CI −37.6 to −34.3) and an increase paid by public insurance (level change: 63.4%, 95% CI 61.8 to 65.0) (Figure 1 D-E, Table 1). By the 2-years post-policy (March 2025), our models estimated 9.3% (95% CI 8.7% to 10.0%) of contraception dispensations in BC were paid out-of-pocket by patients, 82.1% (95% CI 81.1% to 83.2%) by public insurance, and the remaining 8.4% (95% CI 7.5% to 9.3%) by private insurance. Restricting to contraceptives on the BC contraceptive formulary (∼87% of dispensations), at 2-years post-policy, 5.4% (95% CI 4.5% to 6.4%) were paid out-of-pocket (Supplemental Table 2).

**Table 1.**
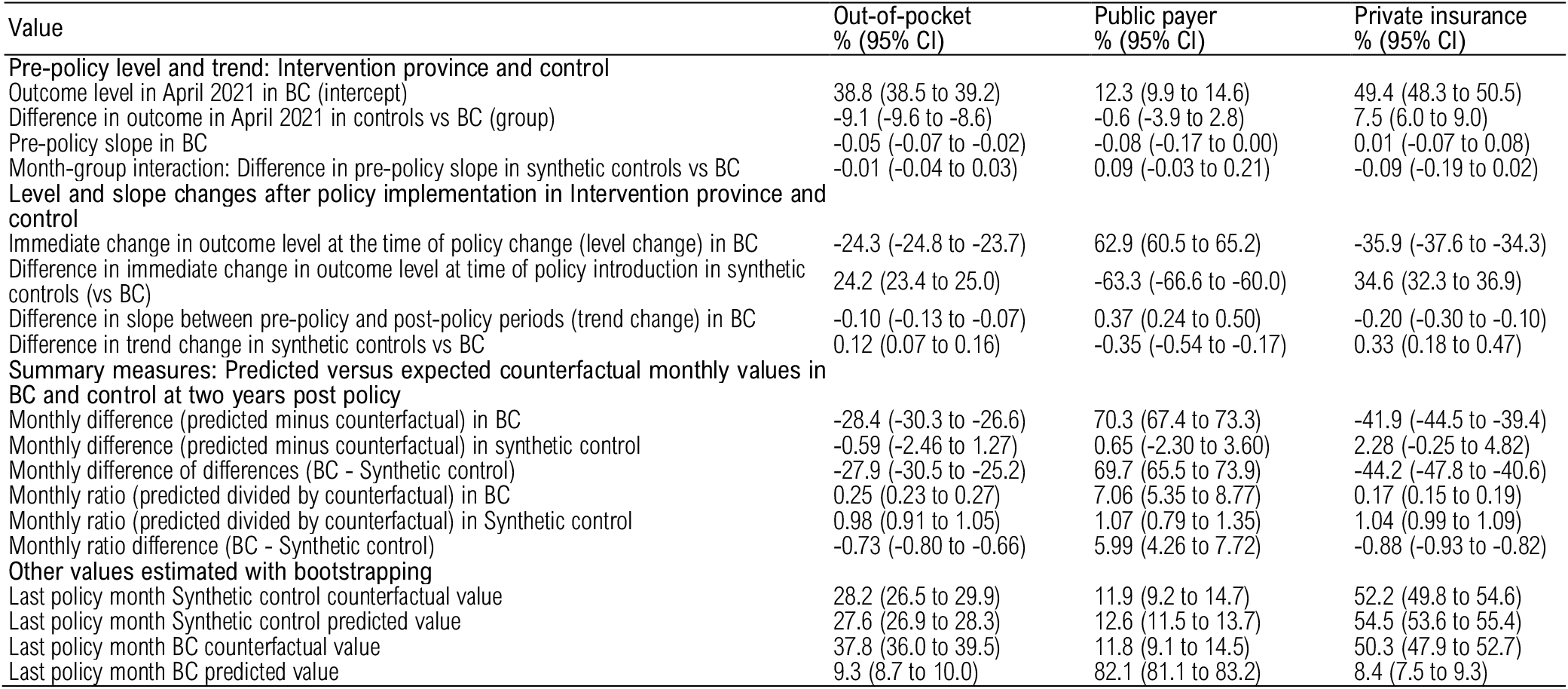
Proportion of contraception dispensations by payer type: CITS model estimates and estimated post-policy values.

| Value | Out-of-pocket<br>% (95% CI) | Public payer<br>% (95% CI) | Private insurance<br>% (95% CI) |
| --- | --- | --- | --- |
| **Pre-policy level and trend: Intervention province and control** |  |  |  |
| Outcome level in April 2021 in BC (intercept) | 38.8 (38.5 to 39.2) | 12.3 (9.9 to 14.6) | 49.4 (48.3 to 50.5) |
| Difference in outcome in April 2021 in controls vs BC (group) | -9.1 (-9.6 to -8.6) | -0.6 (-3.9 to 2.8) | 7.5 (6.0 to 9.0) |
| Pre-policy slope in BC | -0.05 (-0.07 to -0.02) | -0.08 (-0.17 to 0.00) | 0.01 (-0.07 to 0.08) |
| Month-group interaction: Difference in pre-policy slope in synthetic controls vs BC | -0.01 (-0.04 to 0.03) | 0.09 (-0.03 to 0.21) | -0.09 (-0.19 to 0.02) |
| **Level and slope changes after policy implementation in Intervention province and control** |  |  |  |
| Immediate change in outcome level at the time of policy change (level change) in BC | -24.3 (-24.8 to -23.7) | 62.9 (60.5 to 65.2) | -35.9 (-37.6 to -34.3) |
| Difference in immediate change in outcome level at time of policy introduction in synthetic controls (vs BC) | 24.2 (23.4 to 25.0) | -63.3 (-66.6 to -60.0) | 34.6 (32.3 to 36.9) |
| Difference in slope between pre-policy and post-policy periods (trend change) in BC | -0.10 (-0.13 to -0.07) | 0.37 (0.24 to 0.50) | -0.20 (-0.30 to -0.10) |
| Difference in trend change in synthetic controls vs BC | 0.12 (0.07 to 0.16) | -0.35 (-0.54 to -0.17) | 0.33 (0.18 to 0.47) |
| **Summary measures: Predicted versus expected counterfactual monthly values in BC and control at two years post policy** |  |  |  |
| Monthly difference (predicted minus counterfactual) in BC | -28.4 (-30.3 to -26.6) | 70.3 (67.4 to 73.3) | -41.9 (-44.5 to -39.4) |
| Monthly difference (predicted minus counterfactual) in synthetic control | -0.59 (-2.46 to 1.27) | 0.65 (-2.30 to 3.60) | 2.28 (-0.25 to 4.82) |
| Monthly difference of differences (BC - Synthetic control) | -27.9 (-30.5 to -25.2) | 69.7 (65.5 to 73.9) | -44.2 (-47.8 to -40.6) |
| Monthly ratio (predicted divided by counterfactual) in BC | 0.25 (0.23 to 0.27) | 7.06 (5.35 to 8.77) | 0.17 (0.15 to 0.19) |
| Monthly ratio (predicted divided by counterfactual) in Synthetic control | 0.98 (0.91 to 1.05) | 1.07 (0.79 to 1.35) | 1.04 (0.99 to 1.09) |
| Monthly ratio difference (BC - Synthetic control) | -0.73 (-0.80 to -0.66) | 5.99 (4.26 to 7.72) | -0.88 (-0.93 to -0.82) |
| **Other values estimated with bootstrapping** |  |  |  |
| Last policy month Synthetic control counterfactual value | 28.2 (26.5 to 29.9) | 11.9 (9.2 to 14.7) | 52.2 (49.8 to 54.6) |
| Last policy month Synthetic control predicted value | 27.6 (26.9 to 28.3) | 12.6 (11.5 to 13.7) | 54.5 (53.6 to 55.4) |
| Last policy month BC counterfactual value | 37.8 (36.0 to 39.5) | 11.8 (9.1 to 14.5) | 50.3 (47.9 to 52.7) |
| Last policy month BC predicted value | 9.3 (8.7 to 10.0) | 82.1 (81.1 to 83.2) | 8.4 (7.5 to 9.3) |

**Figure 1.**
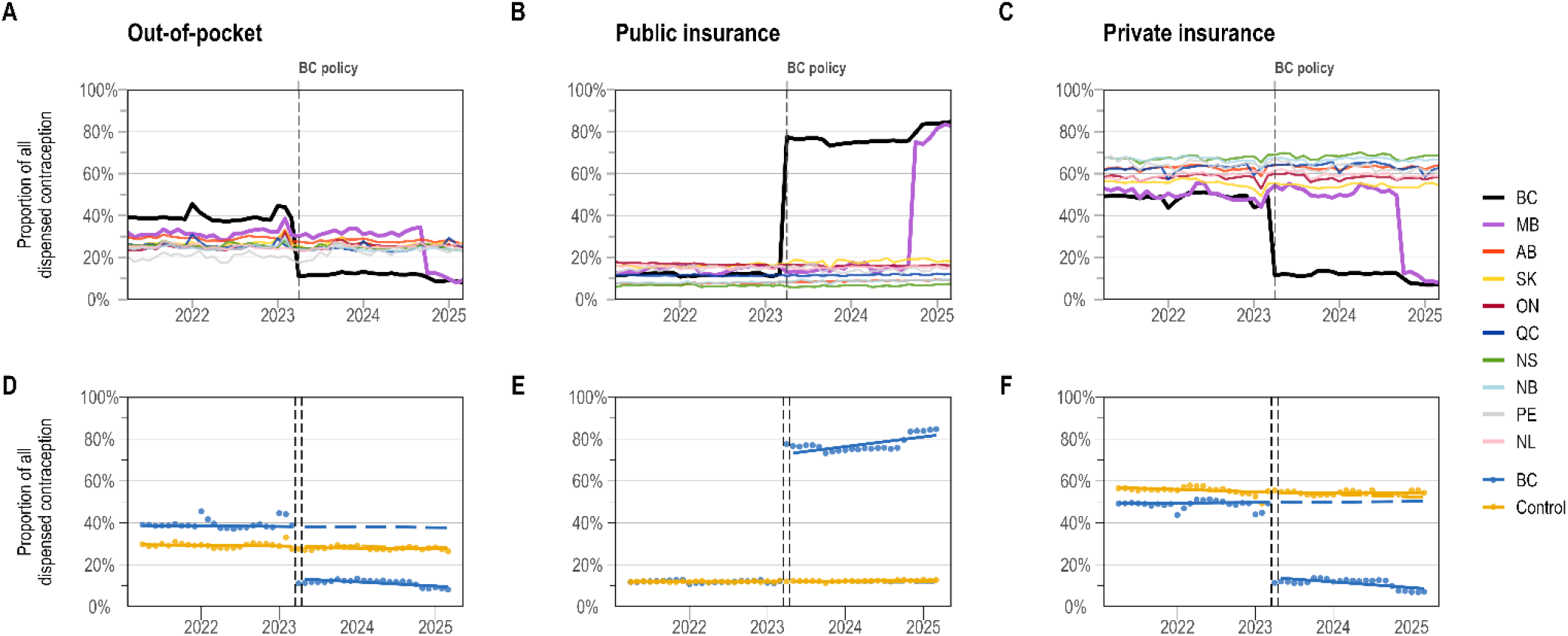
Proportion of dispensed contraception by payer type in all Canadian provinces (A-C) and using a controlled interrupted time series (CITS) for BC and a synthetic control (D-F). *Dashed lines in the CITS graphs show the predicted counterfactual.*

Results were similar in Manitoba with an immediate change in out-of-pocket costs (level change: −20.7%, 95% CI −23.3 to −18.1) (Supplemental Figure 2, Supplemental Table 3) and a decreasing trend (trend change: −0.5%, 95% CI −1.3 to 0.2).

Monthly per capita costs for contraception dispensations in BC decreased immediately for out-of-pocket (level change −$0.59, 95% CI −0.63 to −0.54) and private insurance (−$0.91, 95% CI −0.97 to −0.85) (Figure 2, Table 2). At 2-years post-policy, BC’s estimated out-of-pocket monthly per capita cost was −$0.62 (95% CI −0.72 to −0.53), lower than controls, representing an 86% reduction in out-of-pocket costs compared to control provinces (−0.86, 95% CI −0.96 to −0.75). Among only on-formulary prescriptions, we estimated a 99% reduction in out-of-pocket costs in BC compared to controls (Supplemental Figure 3, Supplemental Table 4), and a 79% reduction in Manitoba (Supplemental Figure 4, Supplemental Table 5).

**Figure 2.**
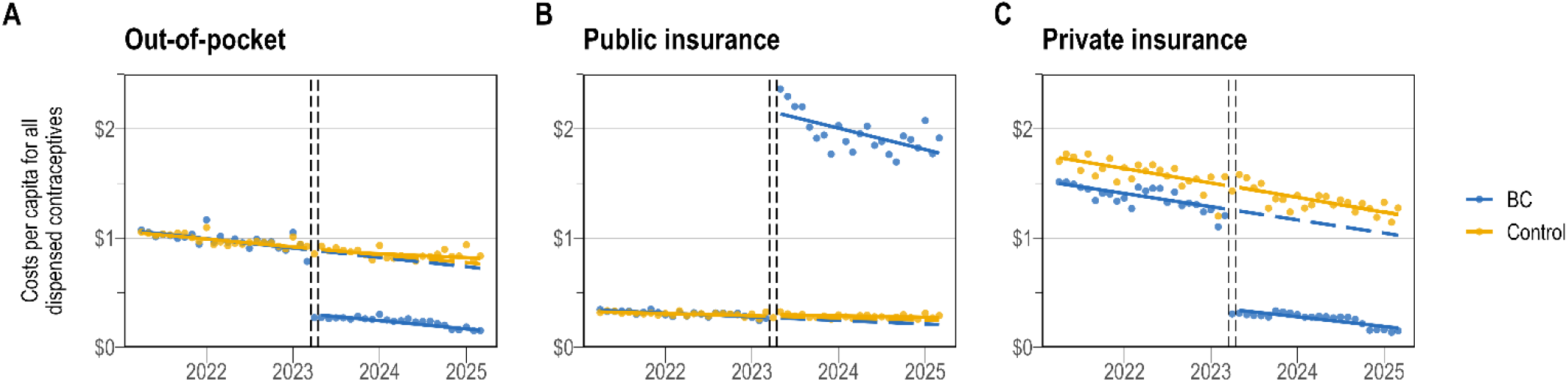
Costs per capita ($/reproductive aged females) of dispensed contraception by payer type using a controlled interrupted time series (CITS) for BC and a synthetic control. *Dashed lines in the CITS graphs show the predicted counterfactual if the policy change had not occurred. Points represent monthly proportions*.

**Table 2.** Costs per capita ($/reproductive aged females) of dispensed contraception by payer type: CITS model estimates and estimated post-policy values.

| Value | Out-of-pocket<br>\$ per capita (95% CI) | Public payer<br>\$ per capita (95% CI) | Private insurance<br>\$ per capita (95% CI) |
| --- | --- | --- | --- |
| **Pre-policy level and trend: Intervention province and control** |  |  |  |
| Outcome level in April 2021 in BC (intercept) | 1.07 (1.04 to 1.10) | 0.40 (0.28 to 0.52) | 1.51 (1.47 to 1.55) |
| Difference in outcome in April 2021 in controls vs BC (group) | -0.02 (-0.06 to 0.03) | -0.07 (-0.25 to 0.10) | 0.24 (0.19 to 0.30) |
| Pre-policy slope in BC | -0.01 (-0.01 to -0.01) | -0.01 (-0.02 to 0.00) | -0.01 (-0.01 to -0.01) |
| Month-group interaction: Difference in pre-policy slope in synthetic controls vs BC | 0.00 (0.00 to 0.00) | 0.01 (0.00 to 0.02) | 0.00 (-0.01 to 0.00) |
| **Level and slope changes after policy implementation in Intervention province and control** |  |  |  |
| Immediate change in outcome level at the time of policy change (level change) in BC | -0.59 (-0.63 to -0.54) | 2.11 (1.98 to 2.23) | -0.91 (-0.97 to -0.85) |
| Difference in immediate change in outcome level at time of policy introduction in synthetic controls (vs BC) | 0.57 (0.51 to 0.63) | -2.09 (-2.27 to -1.91) | 0.92 (0.84 to 1.01) |
| Difference in slope between pre-policy and post-policy periods (trend change) in BC | 0.00 (0.00 to 0.00) | -0.01 (-0.02 to 0.00) | 0.00 (0.00 to 0.01) |
| Difference in trend change in synthetic controls vs BC | 0.00 (0.00 to 0.01) | 0.01 (-0.01 to 0.03) | 0.00 (-0.01 to 0.00) |
| **Summary measures: Predicted versus expected counterfactual monthly values in BC and control at two years post-policy** |  |  |  |
| Monthly difference (predicted minus counterfactual) in BC | -0.57 (-0.64 to -0.50) | 1.55 (1.40 to 1.70) | -0.85 (-0.99 to -0.71) |
| Monthly difference (predicted minus counterfactual) in synthetic control | 0.05 (-0.01 to 0.12) | 0.03 (-0.12 to 0.18) | -0.01 (-0.14 to 0.13) |
| Monthly difference of differences (BC - Synthetic control) | -0.62 (-0.72 to -0.53) | 1.52 (1.31 to 1.74) | -0.85 (-1.04 to -0.65) |
| Monthly ratio (predicted divided by counterfactual) in BC | 0.22 (0.17 to 0.26) | 8.12 (-96.20 to 112.44) | 0.17 (0.11 to 0.23) |
| Monthly ratio (predicted divided by counterfactual) in Synthetic control | 1.07 (0.98 to 1.17) | 1.26 (-0.27 to 2.80) | 1.00 (0.89 to 1.11) |
| Monthly ratio difference (BC RR - Synthetic control RR) | -0.86 (-0.96 to -0.75) | 6.85 (-97.46 to 111.17) | -0.83 (-0.95 to -0.70) |
| **Other values estimated with bootstrapping** |  |  |  |
| Last policy month Synthetic control counterfactual value | 0.76 (0.70 to 0.83) | 0.25 (0.11 to 0.39) | 1.22 (1.09 to 1.34) |
| Last policy month Synthetic control predicted value | 0.82 (0.79 to 0.84) | 0.27 (0.22 to 0.33) | 1.21 (1.16 to 1.26) |
| Last policy month BC counterfactual value | 0.73 (0.66 to 0.79) | 0.21 (0.07 to 0.35) | 1.03 (0.90 to 1.15) |
| Last policy month BC predicted value | 0.16 (0.13 to 0.18) | 1.76 (1.71 to 1.82) | 0.18 (0.12 to 0.23) |

Comparing age groups, we found the largest absolute per capita cost reduction from the BC policy in the 20-29y age group (immediate level change: −$0.95, 95% CI −1.00 to −0.89 and 2-years post-policy change: −$0.80, 95% CI −0.94 to −0.65) (Figure 3, Table 3). Restricting to only on-formulary products did not change the immediate impact but decreased total out-of-pocket costs at 2-years post-policy in all age groups (Supplemental Figure 5, Supplemental Table 6). We observed similar age group patterns for Manitoba (Supplemental Figure 6, Supplemental Table 7).

**Figure 3.**
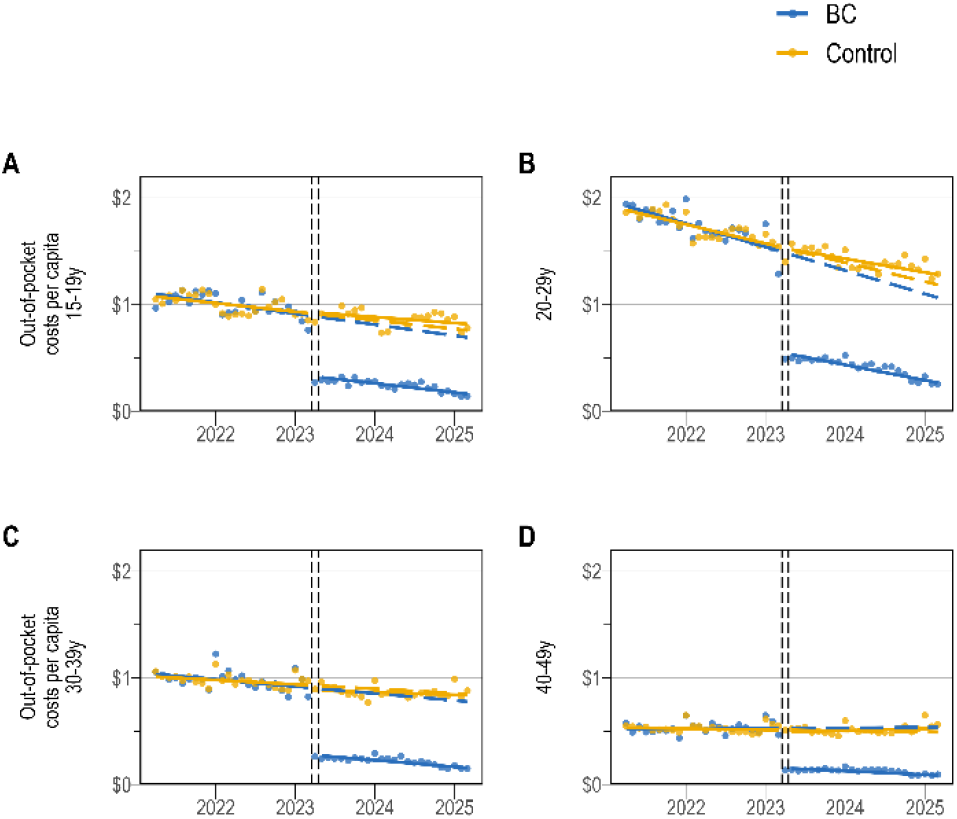
Age-stratified subgroup results for out-of-pocket costs per capita for dispensed contraception using a controlled interrupted time series (CITS) for BC and a synthetic control. *Dashed lines in the CITS graphs show the predicted counterfactual if the policy change had not occurred. Points represent monthly proportions*.

**Table 3.**
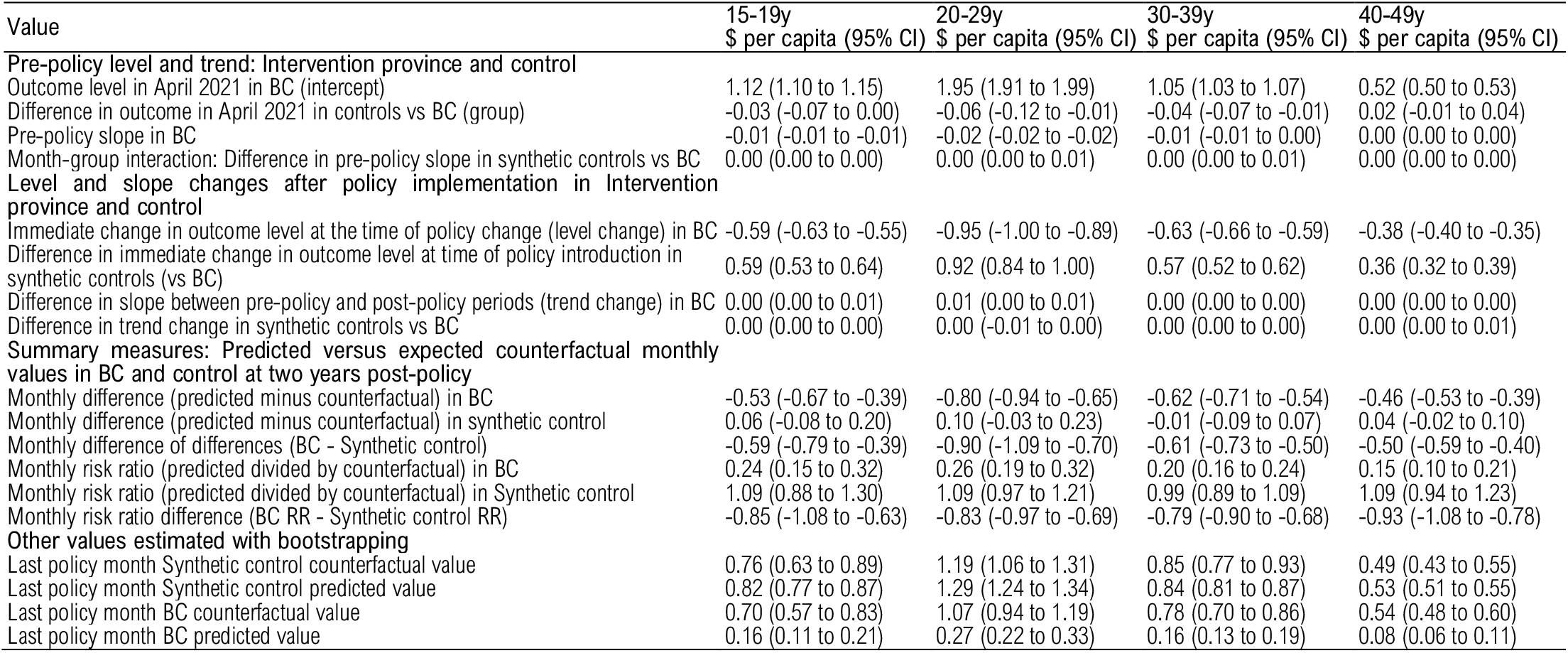
Age-stratified subgroup results for out-of-pocket costs per capita for contraception: CITS model estimates and estimated post-policy values.

| Value | 15-19y<br>\$ per capita (95% CI) | 20-29y<br>\$ per capita (95% CI) | 30-39y<br>\$ per capita (95% CI) | 40-49y<br>\$ per capita (95% CI) |
| --- | --- | --- | --- | --- |
| Pre-policy level and trend: Intervention province and control |  |  |  |  |
| Outcome level in April 2021 in BC (intercept) | 1.12 (1.10 to 1.15) | 1.95 (1.91 to 1.99) | 1.05 (1.03 to 1.07) | 0.52 (0.50 to 0.53) |
| Difference in outcome in April 2021 in controls vs BC (group) | -0.03 (-0.07 to 0.00) | -0.06 (-0.12 to -0.01) | -0.04 (-0.07 to -0.01) | 0.02 (-0.01 to 0.04) |
| Pre-policy slope in BC | -0.01 (-0.01 to -0.01) | -0.02 (-0.02 to -0.02) | -0.01 (-0.01 to 0.00) | 0.00 (0.00 to 0.00) |
| Month-group interaction: Difference in pre-policy slope in synthetic controls vs BC | 0.00 (0.00 to 0.00) | 0.00 (0.00 to 0.01) | 0.00 (0.00 to 0.01) | 0.00 (0.00 to 0.00) |
| Level and slope changes after policy implementation in Intervention province and control |  |  |  |  |
| Immediate change in outcome level at the time of policy change (level change) in BC | -0.59 (-0.63 to -0.55) | -0.95 (-1.00 to -0.89) | -0.63 (-0.66 to -0.59) | -0.38 (-0.40 to -0.35) |
| Difference in immediate change in outcome level at time of policy introduction in synthetic controls (vs BC) | 0.59 (0.53 to 0.64) | 0.92 (0.84 to 1.00) | 0.57 (0.52 to 0.62) | 0.36 (0.32 to 0.39) |
| Difference in slope between pre-policy and post-policy periods (trend change) in BC | 0.00 (0.00 to 0.01) | 0.01 (0.00 to 0.01) | 0.00 (0.00 to 0.00) | 0.00 (0.00 to 0.00) |
| Difference in trend change in synthetic controls vs BC | 0.00 (0.00 to 0.00) | 0.00 (-0.01 to 0.00) | 0.00 (0.00 to 0.00) | 0.00 (0.00 to 0.01) |
| Summary measures: Predicted versus expected counterfactual monthly values in BC and control at two years post-policy |  |  |  |  |
| Monthly difference (predicted minus counterfactual) in BC | -0.53 (-0.67 to -0.39) | -0.80 (-0.94 to -0.65) | -0.62 (-0.71 to -0.54) | -0.46 (-0.53 to -0.39) |
| Monthly difference (predicted minus counterfactual) in synthetic control | 0.06 (-0.08 to 0.20) | 0.10 (-0.03 to 0.23) | -0.01 (-0.09 to 0.07) | 0.04 (-0.02 to 0.10) |
| Monthly difference of differences (BC - Synthetic control) | -0.59 (-0.79 to -0.39) | -0.90 (-1.09 to -0.70) | -0.61 (-0.73 to -0.50) | -0.50 (-0.59 to -0.40) |
| Monthly risk ratio (predicted divided by counterfactual) in BC | 0.24 (0.15 to 0.32) | 0.26 (0.19 to 0.32) | 0.20 (0.16 to 0.24) | 0.15 (0.10 to 0.21) |
| Monthly risk ratio (predicted divided by counterfactual) in Synthetic control | 1.09 (0.88 to 1.30) | 1.09 (0.97 to 1.21) | 0.99 (0.89 to 1.09) | 1.09 (0.94 to 1.23) |
| Monthly risk ratio difference (BC RR - Synthetic control RR) | -0.85 (-1.08 to -0.63) | -0.83 (-0.97 to -0.69) | -0.79 (-0.90 to -0.68) | -0.93 (-1.08 to -0.78) |
| Other values estimated with bootstrapping |  |  |  |  |
| Last policy month Synthetic control counterfactual value | 0.76 (0.63 to 0.89) | 1.19 (1.06 to 1.31) | 0.85 (0.77 to 0.93) | 0.49 (0.43 to 0.55) |
| Last policy month Synthetic control predicted value | 0.82 (0.77 to 0.87) | 1.29 (1.24 to 1.34) | 0.84 (0.81 to 0.87) | 0.53 (0.51 to 0.55) |
| Last policy month BC counterfactual value | 0.70 (0.57 to 0.83) | 1.07 (0.94 to 1.19) | 0.78 (0.70 to 0.86) | 0.54 (0.48 to 0.60) |
| Last policy month BC predicted value | 0.16 (0.11 to 0.21) | 0.27 (0.22 to 0.33) | 0.16 (0.13 to 0.19) | 0.08 (0.06 to 0.11) |

Per capita costs for all prescription contraception (combined dispensation fees and direct drug/device costs) were decreasing in both BC and controls in the pre-policy period (Figure 4). Following the BC policy change, monthly per capita costs immediately increased (level change: $0.29, 95% CI 0.26 to 0.32) relative to the counterfactual and the decreasing cost trend showed a small increase (trend change: −$0.01, 95% CI −0.01 to −0.01) (Table 4). At 2-years post-policy, monthly costs in BC compared to control provinces were unchanged for all contraception ($0.10, 95% CI −0.24 to 0.44), higher for LARC ($0.36, 95% CI 0.18 to 0.54) and lower for SARC (−$0.22, 95% CI −0.42 to 0.03) (Supplemental Table 8). At 2-years post-policy, the absolute monthly per-capita pharmacy-related costs for contraception dispensations in BC was $2.12 (95% CI 2.03 to 2.21). Using ITS models for total costs, (Supplemental Table 8, Supplemental Figure 7) the increased cost for prescription contraceptives with BC’s public insurance as primary payer was $30.5 million (95% CI $29.0 to $31.9 million) in year 1 (April 1, 2023 to March 30, 2023) and $28.5 million (95% CI $26.4 to $30.6 million) in year 2 (April 1, 2024 to March 30, 2025).

**Figure 4.**
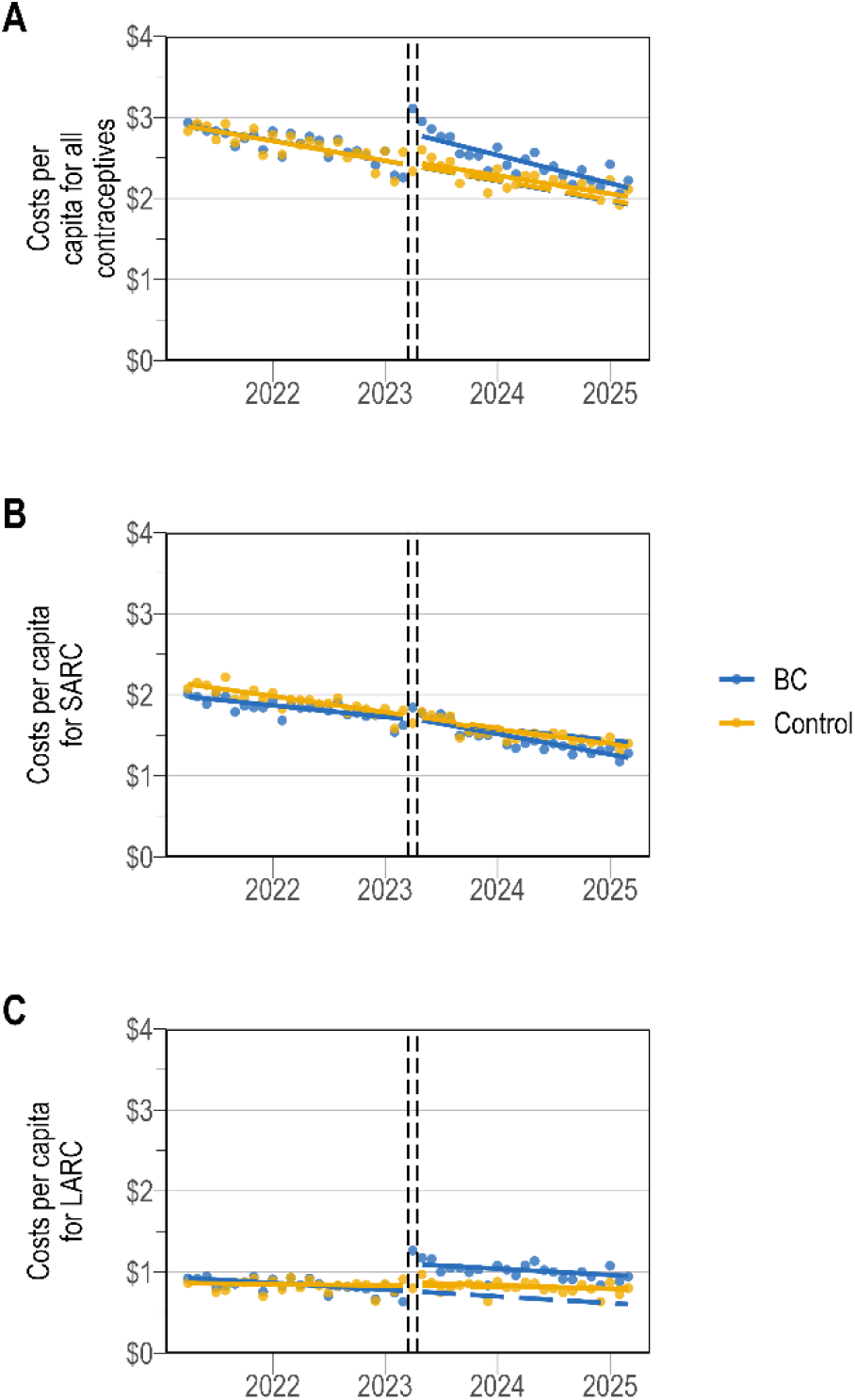
Total costs per capita ($/reproductive aged females) for all dispensed contraception, SARC and LARC using a controlled interrupted time series (CITS) for BC and a synthetic control. *Dashed lines in the CITS graphs show the predicted counterfactual if the policy change had not occurred. Points represent monthly proportions*.

**Table 4.** Contraception costs per capita ($/reproductive aged females) for all contraception, SARC and LARC: CITS model estimates and estimated post-policy values.

| Value | All costs per capita<br>\$ (95% CI) | SARC costs per capita<br>\$ (95% CI) | LARC costs per capita<br>\$ (95% CI) |
| --- | --- | --- | --- |
| **Pre-policy level and trend: Intervention province and control** |  |  |  |
| Outcome level in April 2021 in BC (intercept) | 2.92 (2.90 to 2.94) | 2.00 (1.98 to 2.02) | 0.94 (0.93 to 0.95) |
| Difference in outcome in April 2021 in controls vs BC (group) | 0.01 (-0.02 to 0.04) | 0.16 (0.13 to 0.20) | -0.07 (-0.08 to -0.05) |
| Pre-policy slope in BC | -0.02 (-0.02 to -0.02) | -0.01 (-0.01 to -0.01) | -0.01 (-0.01 to -0.01) |
| Month-group interaction: Difference in pre-policy slope in synthetic controls vs BC | 0.00 (0.00 to 0.00) | 0.00 (-0.01 to 0.00) | 0.00 (0.00 to 0.01) |
| **Level and slope changes after policy implementation in Intervention province and control** |  |  |  |
| Immediate change in outcome level at the time of policy change (level change) in BC | 0.29 (0.26 to 0.32) | -0.04 (-0.07 to 0.00) | 0.31 (0.30 to 0.33) |
| Difference in immediate change in outcome level at time of policy introduction in synthetic controls (vs BC) | -0.33 (-0.38 to -0.28) | -0.05 (-0.10 to 0.00) | -0.27 (-0.29 to -0.26) |
| Difference in slope between pre-policy and post-policy periods (trend change) in BC | -0.01 (-0.01 to -0.01) | -0.01 (-0.01 to -0.01) | 0.00 (0.00 to 0.00) |
| Difference in trend change in synthetic controls vs BC | 0.01 (0.01 to 0.02) | 0.01 (0.01 to 0.01) | 0.00 (0.00 to 0.00) |
| **Summary measures: Predicted versus expected counterfactual monthly values in BC and control at two years post policy** |  |  |  |
| Monthly difference (predicted minus counterfactual) in BC | 0.19 (-0.05 to 0.43) | -0.21 (-0.35 to -0.06) | 0.35 (0.22 to 0.48) |
| Monthly difference (predicted minus counterfactual) in synthetic control | 0.09 (-0.15 to 0.33) | 0.02 (-0.12 to 0.16) | -0.02 (-0.14 to 0.11) |
| Monthly difference of differences (BC - Synthetic control) | 0.10 (-0.24 to 0.44) | -0.23 (-0.43 to -0.03) | 0.36 (0.18 to 0.54) |
| Monthly ratio (predicted divided by counterfactual) in BC | 1.10 (0.96 to 1.23) | 0.86 (0.77 to 0.95) | 1.58 (1.26 to 1.91) |
| Monthly ratio (predicted divided by counterfactual) in Synthetic control | 1.05 (0.92 to 1.18) | 1.02 (0.91 to 1.12) | 0.98 (0.83 to 1.14) |
| Monthly ratio difference (BC - Synthetic control) | 0.05 (-0.14 to 0.24) | -0.16 (-0.29 to -0.02) | 0.60 (0.24 to 0.96) |
| **Other values estimated with bootstrapping** |  |  |  |
| Last policy month Synthetic control counterfactual value | 1.96 (1.74 to 2.19) | 1.38 (1.25 to 1.51) | 0.80 (0.69 to 0.92) |
| Last policy month Synthetic control predicted value | 2.05 (1.96 to 2.14) | 1.40 (1.35 to 1.45) | 0.79 (0.74 to 0.83) |
| Last policy month BC counterfactual value | 1.96 (1.74 to 2.18) | 1.44 (1.32 to 1.57) | 0.61 (0.49 to 0.72) |
| Last policy month BC predicted value | 2.15 (2.06 to 2.23) | 1.24 (1.18 to 1.29) | 0.95 (0.90 to 1.01) |

## DISCUSSION

BC’s contraception policy (April 1, 2023) substantially reduced the proportion of contraceptives paid out-of-pocket from 39% (pre-policy) to 11% at 2-years post-policy (6% for contraceptives on formulary). Monthly out-of-pocket costs for contraception also decreased immediately, resulting in a 76% reduction in out-of-pocket costs at 2-years post-policy compared to expected trends. Following the policy change, overall per capita costs in BC increased because of increased LARC dispensations while SARC costs decreased. Pre-policy, out-of-pocket costs were highest in the 20-29y age group and we observed the largest immediate impact of policy on decreasing costs in this group. Despite the limited post-policy interval, results for Manitoba were generally consistent following a similar policy.

While we reported costs per capita among all reproductive aged females, not all females are prescription contraception users. In a previous analysis, we found that 23% of BC reproductive-aged females used prescription contraception in 2021 ^42^, similar to other countries ^54^, although fewer than 30% estimated in a 2021 national survey ^55^. If we apply our results to the proportion of females who are contraception users, we estimate an out-of-pocket cost reduction of approximately $30 per user (ages 15-49y) per year. For youth ages 15 to 19y who have the lowest rate of contraception use (within their age bracket), at 2-years post policy we estimate a yearly cost reduction of more than $50 per contraceptive user. Given that long-acting methods cost approximately $400 over 5–7 years, whereas short-acting methods cost about $30 per month, the real-world cost savings per person would differ according to the contraceptive method used.

Our finding that almost 40% of *contraception* costs were paid out-of-pocket in BC pre-policy is substantially different than the 20% of *all* prescription drugs paid out-of-pocket in Canada ^30^. Further, using our synthetic control as a weighted average of all provinces except BC and Manitoba, we found that 30% of costs were paid out-of-pocket for contraception in other provinces. There are several possible reasons for this. First, contraception users, especially youth, express a strong need for privacy ^26^ and may choose to pay out-of-pocket to maintain confidentiality when sharing a private insurance plan with a parent or partner. Second, even if users are covered by a private or public insurance plan, patients in BC pay out-of-pocket until they reach their annual deductible. Reproductive-aged individuals generally have lower drug expenditures, so many will never reach their deductible and thus pay entirely out-of-pocket. Comparing BC to other provinces pre-policy, we found both the highest proportion of contraception paid out-of-pocket and the lowest proportion paid by private insurance. While reasons for this are unclear, this aligns with the 2019 Canadian Community Health Survey (CCHS) data showing that both BC and Manitoba had the highest proportion of females, across all age groups, without any drug plan coverage ^23^.

The effect of BC’s policy on out-of-pocket costs was larger ^56–58^ or comparable to ^59^ in the United States following the Affordable Care Act ^57,58^. BC’s free contraception policy also changed contraceptive method-mix, in particular with an increase in LARC use, primarily driven by new users and those that switched from SARC to LARC ^42^. Previously in Canada, cost-related medication nonadherence was more likely for females than males, even after controlling for drug insurance coverage ^23,60^. The cost impacts of a universal contraception policy, combined with the impacts on method choice and uptake, suggests 1) there was unmet need overall for contraception and 2) choice of method and uptake were clearly impacted by cost barriers.

This study provides a cost-based analysis of universal, first-dollar contraception coverage policies in two Canadian provinces with distinct and demographically diverse populations. We used a national-level prescription dispensation dataset that allows for cross-province comparisons, a robust quasi-experimental study design ^35,37^, and a synthetic control derived from other Canadian provinces. Our study should be interpreted with some limitations. We report costs for prescriptions dispensed at the pharmacy and included an average dispensation fee without other costs that could be incurred by the patient or the health system (i.e., pharmacy markups, consultation fees for pharmacy prescribing, health care provider insertion costs for IUDs or implants). There were other cost-related changes during the follow-up period. Specifically, in September 2023 a new generic for the most common oral contraceptive came on the market leading to a price drop for this group ^61^; however, this was consistent in all provinces so we believe this had no meaningful effect on our analysis. While new products were added to the formulary, we did not model this separately because formulary modifications are an expected feature of policy changes. This evaluation did not include an overall assessment of changes in reproductive health service costs (e.g., costs averted through preventing unintended pregnancy, permanent contraception procedures, or hysterectomy to manage idiopathic peri-/post-menopausal bleeding), nor were we able to examine measures of equity or poor access other than age. These remain areas for future research we will explore when sufficient post-policy data has accrued to measure these longer-term impacts or with other data sources.

BC’s universal, first-dollar coverage for prescription contraceptives resulted in a substantial decrease in out-of-pocket costs to all contraceptive users and decreased private insurance costs. Youth contraceptive users previously had the highest out-of-pocket costs and also the greatest cost reductions because of this policy. Universal, first-dollar coverage is highly effective at lowering the out-of-pocket cost burden of prescription contraception use.

## Data Availability

Data used in this study was obtained under license from IQVIA Solutions Canada Inc and cannot be shared by the authors. The statements, findings, conclusions, views, and opinions expressed in this report are based in part on data obtained under license from IQVIA Solutions Canada Inc. concerning the following information service(s): GPM, from: September 1st, 2016 to March 31st, 2025. All Rights Reserved. The statements, findings, conclusions, views, and opinions expressed herein are not necessarily those of IQVIA Inc. or any of its affiliated or subsidiary entities.

## Data disclaimer

The statements, findings, conclusions, views, and opinions expressed in this report are based in part on data obtained under license from IQVIA Solutions Canada Inc. concerning the following information service(s): GPM, from: September 1^st^, 2016 to March 31st, 2025. All Rights Reserved. The statements, findings, conclusions, views, and opinions expressed herein are not necessarily those of IQVIA Inc. or any of its affiliated or subsidiary entities.

