## Supplemental Materials for "Impacts of British Columbia’s free contraception policy on out-of-pocket payments and contraception costs: an interrupted time series analysis with other provinces as controls"

#### SUPPLEMENTAL TABLES & FIGURES

##### Supplemental Table 1. Product descriptions, DINs and annual costs per unit

| Type | Product | DIN | Manufacturer | BC formulary coverage | $/unit BC 2022 | $/unit BC 2024-5 | $/unit ON 2022 |
|---|---|---|---|---|---|---|---|
| DMPA | DEPO-PROVERA SYRINGE 150MG/ML 1ML #US376119 | 2523493 | PFIZER | Full benefit | 31.86 | 32.39 | 31.45 |
| DMPA | DEPO-PROVERA VIAL 150MG/ML 1ML | 585092 | PFIZER | Full benefit | 31.66 | 32.15 | 31.27 |
| DMPA | MEDROXYPROGESTERONE VIAL 150MG/ML 1ML #10390 | 2322250 | SANDOZ CANADA INC | Full benefit | 20.86 | | |
| IMPLANT | NEXPLANON IMPLANT 68MG 1 #1042025 | 2499509 | ORGANON | Full benefit | 299.94 | 293.51 | 294.84 |
| IUD | GYNE T I.U.D. VARIOUS | 306460 | JANSSEN PHARMA | Full benefit | 0.00 | 0.00 | |
| IUD | JAYDESS IUD 13.5MG 1 UNIT | 2408295 | BAYER HEALTHCARE | not covered | 278.26 | 277.73 | 277.73 |
| IUD | KYLEENA IUD 19.5MG 1 #5650093257 | 2459523 | BAYER HEALTHCARE | Full benefit | 338.16 | 350.42 | 334.76 |
| IUD | MIRENA I.U.D. 52MG 1 (INTRAUTERINE SYSTEM) | 2243005 | BAYER HEALTHCARE | Full benefit | 345.89 | 372.82 | 345.44 |
| OCP | ALESSE TAB 100MCG/20MCG (21) 21 | 2236974 | PFIZER | Partial benefit | 0.73 | 0.73 | 0.73 |
| OCP | ALESSE TAB 100MCG/20MCG (28) 28 | 2236975 | PFIZER | Partial benefit | 0.55 | 0.55 | 0.55 |
| OCP | ALYSENA 21 TAB 100MCG/20MCG (21) 7X3 BL ST(21) | 2387875 | APOTEX INC | Full benefit | 0.40 | 0.20 | 0.40 |
| OCP | ALYSENA 28 TAB 100MCG/20MCG (28) 7X4 BL ST(28) | 2387883 | APOTEX INC | Full benefit | 0.30 | 0.15 | 0.30 |
| OCP | APRI TAB 150MCG/30MCG (21) 21 #50512 | 2317192 | TEVA CANADA LTD | Full benefit | 0.40 | 0.40 | 0.40 |
| OCP | APRI TAB 150MCG/30MCG (28) 28 #50510 | 2317206 | TEVA CANADA LTD | Full benefit | 0.30 | 0.30 | 0.30 |
| OCP | AUDRINA 21 TAB 100MCG/20MCG 21X1 BL ST(21) | 2532174 | JAMP PHARMA | Full benefit | | 0.29 | |
| OCP | AUDRINA 28 TAB 100MCG/20MCG 28X1 BL ST(28) | 2532182 | JAMP PHARMA | Full benefit | | 0.22 | |
| OCP | AVIANE TAB 100MCG/20MCG (21) 21 | 2298538 | TEVA CANADA LTD | Full benefit | 0.40 | 0.20 | 0.40 |
| OCP | AVIANE TAB 100MCG/20MCG (28) 28 | 2298546 | TEVA CANADA LTD | Full benefit | 0.30 | 0.15 | 0.30 |
| OCP | BREVICON 0.5/35 TAB 0.5MG/35MCG (21) 21 | 00373265;02187086 | PFIZER | Full benefit | 0.69 | 0.69 | 0.69 |
| OCP | BREVICON 0.5/35 TAB 0.5MG/35MCG (28) 28 | 00373273;02187094 | PFIZER | Full benefit | 0.52 | 0.52 | 0.52 |
| OCP | BREVICON 1/35 TAB 1MG/35MCG (21) 21X10 | 00531006;02162563;02189054 | PFIZER | Full benefit | 0.69 | 0.69 | 0.69 |
| OCP | BREVICON 1/35 TAB 1MG/35MCG (28) 28X10 | 00531014;02189062 | PFIZER | Full benefit | 0.52 | 0.52 | 0.52 |
| OCP | CYCLEN TAB 250MCG/35MCG (21) 21 | 01968440;8942 | JANSSEN PHARMA | not covered | 1.28 | | 1.28 |
| OCP | CYCLEN TAB 250MCG/35MCG (28) 28 | 1992872 | JANSSEN PHARMA | not covered | 0.96 | 0.96 | 0.96 |
| OCP | DEMULEN 30 TAB 2MG/30MCG (21) 21X10 COMPACK | 469327 | PFIZER | not covered | 0.73 | | |
| OCP | DEMULEN 30 TAB 2MG/30MCG (28) 28X5 COMPACK | 471526 | PFIZER | not covered | | | 0.58 |
| OCP | DROSPIRENONE&ET ESTRA 21 FC TAB 3MG/30MCG (21) 21X1 BL ST(21) | 2421437 | GLENMARK PHARMA CANA | Full benefit | | 0.30 | |
| OCP | DROSPIRENONE&ET ESTRA 28 FC TAB 3MG/30MCG (28) 28X1 BL ST(28) | 2421445 | GLENMARK PHARMA CANA | Full benefit | | 0.22 | 0.23 |
| OCP | DROSPIRENONE&ETHINL STRA FC TAB 3MG/20MCG 28X3 BL ST(84) | 2462060 | GLENMARK PHARMA CANA | Full benefit | | 0.38 | 0.30 |
| OCP | ESME TAB 100MCG/20MCG (21) 21 | 2388138 | MYLAN PHARMA | not covered | | | 0.41 |
| OCP | FREYA TAB 150MCG/30MCG (21) 21 | 2396491 | MYLAN PHARMA | Full benefit | 0.41 | 0.41 | 0.41 |
| OCP | FREYA TAB 150MCG/30MCG (28) 28 | 2396610 | MYLAN PHARMA | Full benefit | 0.31 | 0.31 | 0.31 |
| OCP | INDAYO TAB 150MCG/30MCG 91#400523783 | 2398869 | MYLAN PHARMA | not covered | 0.51 | 0.51 | 0.51 |
| OCP | JENCYCLA TAB 350MCG 28X1 BL ST(28) #44071698 | 2441306 | LUPIN PHARMA CDN LTD | Full benefit | 0.39 | 0.39 | 0.39 |
| OCP | LINESSA TAB 150MCG/125MCG (21) 21 | 2272903 | ASPEN PHARMA TRADING | Full benefit | 0.79 | 0.86 | 0.80 |
| OCP | LINESSA TAB 150MCG/125MCG (28) 28 | 2257238 | ASPEN PHARMA TRADING | Full benefit | 0.59 | 0.65 | 0.59 |
| OCP | LOESTRIN 1.5/30 TAB 1.5MG/30MCG (21) 21 | 297143 | ALLERGAN | not covered | 0.75 | 0.75 | 0.75 |
| OCP | LOESTRIN 1.5/30 TAB 1.5MG/30MCG (28) 28 | 353027 | ALLERGAN | not covered | 0.56 | 0.56 | 0.56 |
| OCP | LOLO TAB 1MG/10MCG/10MCG 28X6 BL ST(168)[1] | 2417456 | ABBVIE | not covered until Oct 14, 2025, Full benefit (as of Oct 15, 2025) | 0.64 | 0.67 | 0.64 |
| OCP | MARVELON TAB 150MCG/30MCG (21) 21 #1036166 | 2042487 | ORGANON | Partial benefit | 0.87 | 0.89 | 0.86 |
| OCP | MARVELON TAB 150MCG/30MCG (28) 28 | 2042479 | ORGANON | Partial benefit | 0.65 | 0.67 | 0.65 |
| OCP | MICRONOR TAB 350MCG 28X6 DIALPACK | 37605 | JANSSEN PHARMA | not covered | 0.96 | 0.96 | 0.96 |
| OCP | MIN-OVRAL TAB 150MCG/30MCG (21) 21X12 SLEEVE | 00782432;02042320 | PFIZER | Partial benefit | 0.82 | 0.88 | 0.82 |
| OCP | MIN-OVRAL TAB 150MCG/30MCG (28) 28X12 SLEEVE | 00782440;02042339 | PFIZER | Partial benefit | 0.62 | 0.66 | 0.62 |
| OCP | MINESTRIN 1/20 TAB 1MG/20MCG (21) 21 | 315966 | ALLERGAN | not covered | 0.75 | | 0.75 |
| OCP | MINESTRIN 1/20 TAB 1MG/20MCG (28) 28 | 343838 | ALLERGAN | not covered | 0.56 | 0.57 | 0.56 |
| OCP | MIRVALA 21 TAB 150MCG/30MCG (21) 7X3 BL ST (21) | 2410249 | APOTEX INC | Full benefit | 0.40 | 0.40 | 0.40 |
| OCP | MIRVALA 28 TAB 150MCG/30MCG (28) 7X4 BL ST (28) | 2410257 | APOTEX INC | Full benefit | 0.30 | 0.30 | 0.30 |
| OCP | MOVISSE TAB 350MCG 28X1 BL ST(28) #400522289 | 2410303 | MYLAN PHARMA | Full benefit | 0.39 | 0.39 | 0.39 |
| OCP | MYA TAB 3MG/20MCG 28X1 BL ST(28) | 2415380 | APOTEX INC | Full benefit | 0.42 | 0.38 | 0.43 |
| OCP | NEXTSTELLIS FC TAB 15MG/3MG 28X1 BL ST(28) | 2513218 | SEARCHLIGHT PHARMA | not covered | 0.63 | 0.63 | 0.63 |
| OCP | ORTHO-CEPT TAB 150MCG/30MCG (28) 28 COMPACT | 2042533 | JANSSEN PHARMA | not covered | 0.75 | | |
| OCP | ORTHO 1/35 TAB 1MG/35MCG (21) 21X6 | 372846 | JANSSEN PHARMA | not covered | | | 1.10 |
| OCP | OVIMA TAB 150MCG/30MCG (21) 21 | 2387085 | APOTEX INC | Full benefit | 0.41 | 0.41 | 0.41 |
| OCP | OVIMA TAB 150MCG/30MCG (28) 28 | 2387093 | APOTEX INC | Full benefit | 0.31 | 0.31 | 0.31 |
| OCP | PORTIA TAB 150MCG/30MCG (21) 21 | 2295946 | TEVA CANADA LTD | Full benefit | 0.41 | 0.41 | 0.41 |
| OCP | PORTIA TAB 150MCG/30MCG (28) 28 | 2295954 | TEVA CANADA LTD | Full benefit | 0.31 | 0.31 | 0.31 |
| OCP | SEASONALE FC TAB 150MCG/30MCG 91 #84001-0190 | 2296659 | TEVA WOMENS HEALTH | not covered | 0.72 | 0.83 | 0.72 |
| OCP | SEASONIQUE FC TAB 150MCG/30MCG/10MCG 91 #84000-0190 | 2346176 | TEVA WOMENS HEALTH | not covered | 0.67 | 0.76 | 0.67 |
| OCP | SELECT 1/35 TAB 1MG/35MCG (21) 21 | 2197502 | PFIZER | not covered | 0.51 | 0.52 | 0.51 |
| OCP | SELECT 1/35 TAB 1MG/35MCG (28) 28 | 2199297 | PFIZER | not covered | 0.38 | 0.39 | 0.38 |

[1] Lolo was the 3rd most commonly dispensed OCP in BC in 2022-2024, ~8-9% of OCP, and was added to the BC formulary on October 15, 2024.

| Type | Product | DIN | Manufacturer | BC formulary coverage | $/unit BC 2022 | $/unit BC 2024-5 | $/unit ON 2022 |
|---|---|---|---|---|---|---|---|
| OCP | SLYND FC TAB 4MG 28X1 BL ST(28) #PF-0380[2] | 2522802 | DUCHESNAY LABS | Not covered to Dec 9, 2025, Full benefit (Dec 10, 2025) | 0.45 | 0.45 | 0.45 |
| OCP | SYNPHASIC TAB 1MG/0.5MG/35MCG (21) 21X10 | 00620947;02187108 | PFIZER | Full benefit | 0.62 | 0.63 | 0.62 |
| OCP | SYNPHASIC TAB 1MG/0.5MG/35MCG (28) 28X10 | 00695734;02187116 | PFIZER | Full benefit | 0.47 | 0.47 | 0.47 |
| OCP | SYNPHASIC TAB 1MG/0.5MG/35MCG (28) 28X10 #2710705 | | PFIZER | not covered | | 0.49 | |
| OCP | TRI-CIRA 21 TAB 250MCG/215MCG (21) 21X1 BL ST (21) #64523 | 2508087 | APOTEX INC | Full benefit | 0.67 | 0.68 | 0.68 |
| OCP | TRI-CIRA 28 TAB 250MCG/215MCG (28) 28X1 BL ST (28) #64524 | 2508095 | APOTEX INC | Full benefit | 0.51 | 0.51 | 0.51 |
| OCP | TRI-CYCLEN LO TAB 250MCG/215MCG (21) 21 BLISTER | 2258560 | JANSSEN PHARMA | not covered | 0.87 | | 0.87 |
| OCP | TRI-CYCLEN LO TAB 250MCG/215MCG (28) 28 BLISTER | 2258587 | JANSSEN PHARMA | not covered | 0.66 | | 0.64 |
| OCP | TRI-CYCLEN TAB 250MCG/215MCG (21) 21 COMPACT | 02028700;02229218 | JANSSEN PHARMA | not covered | | | 1.28 |
| OCP | TRI-CYCLEN TAB 250MCG/215MCG (28) 28 COMPACT | 02029421;02229226 | JANSSEN PHARMA | not covered | 0.96 | | 0.96 |
| OCP | TRI-JORDYNA 21 TAB 250MCG/215MCG (21) 21X3 #5013230364 | 2486296 | GLENMARK PHARMA CANA | Full benefit | 0.68 | 0.68 | 0.68 |
| OCP | TRI-JORDYNA 28 TAB 250MCG/215MCG (28) 28 | 2486318 | GLENMARK PHARMA CANA | Full benefit | 0.51 | 0.51 | 0.51 |
| OCP | TRICIRA LO 21 DAY TAB 250MCG/215MCG (21) 21 BL ST(21) | 2401967 | APOTEX INC | Full benefit | 0.65 | 0.70 | 0.65 |
| OCP | TRICIRA LO 28 DAY TAB 250MCG/215MCG (28) 28 BL ST(28) | 2401975 | APOTEX INC | Full benefit | 0.48 | 0.52 | 0.49 |
| OCP | TRIQUILAR TAB 125MCG/75MCG/50MCG (21) 21 | 707600 | BAYER HEALTHCARE | Full benefit | 0.74 | 0.74 | 0.74 |
| OCP | TRIQUILAR TAB 125MCG/75MCG/50MCG (28) 28 | 707503 | BAYER HEALTHCARE | Full benefit | 0.56 | 0.56 | 0.56 |
| OCP | YASMIN FC TAB 3MG/30MCG (21) 21 | 2261723 | BAYER HEALTHCARE | Partial benefit | 0.59 | 0.59 | 0.59 |
| OCP | YASMIN FC TAB 3MG/30MCG (28) 28 | 2261731 | BAYER HEALTHCARE | Partial benefit | 0.44 | 0.44 | 0.44 |
| OCP | YAZ FC TAB 3MG/20MCG 28 | 2321157 | BAYER HEALTHCARE | Partial benefit | 0.57 | 0.57 | 0.57 |
| OCP | YAZ PLUS FC TAB 3MG/451MCG/451MCG/20MCG 28X1 BL ST(28) | 2387433 | BAYER HEALTHCARE | not covered | 0.42 | 0.42 | 0.42 |
| OCP | ZAMINE 21 FC TAB 3MG/30MCG (21) 7X3 BL ST(21) #62360 | 2410788 | APOTEX INC | Full benefit | 0.38 | 0.30 | 0.38 |
| OCP | ZAMINE 28 FC TAB 3MG/30MCG (28) 7X4 BL ST(28) #62361 | 2410796 | APOTEX INC | Full benefit | 0.30 | 0.23 | 0.30 |
| PATCH[3] | EVRA PATCH 200MCG/35MCG 1 #61052 | 02246340;02248297 | SEARCHLIGHT PHARMA | not covered | 7.75 | 8.20 | 7.73 |
| RING | HALOETTE SR RING 11.7MG/2.7MG 1 SACHET #B0006010 | 2520028 | SEARCHLIGHT PHARMA | Full benefit | 13.53 | 13.14 | 13.37 |
| RING | NUVARING SR RING 11.4MG/2.6MG 1 | 2253186 | ORGANON | Partial benefit | 17.30 | 18.19 | 17.35 |

[2] Slynd was the 11th most common OCP in BC, from 0% in 2022 to 2.5% in March 2025 and was added to the BC formulary on December 10, 2024.

[3] The patch is not covered on the BC contraceptives formulary as of June 30, 2025.

**Supplemental Table 2. Proportions by payer type for contraceptives on BC formulary: CITS model estimates and estimated post-policy values**

| Value | Out-of-pocket % (95%CI) | Public payer % (95%CI) | Private insurance % (95%CI) |
|---|---|---|---|
| Pre-policy level and trend: Intervention province and control | | | |
| Outcome level in April 2021 in BC (intercept) | 38.1 (36.8 to 39.4) | 13.5 (12.6 to 14.3) | 48.4 (47.4 to 49.3) |
| Difference in outcome in April 2021 in controls vs BC (group) | -9.4 (-11.2 to -7.5) | 0.0 (-1.2 to 1.1) | 8.9 (7.6 to 10.3) |
| Pre-policy slope in BC | 0.0 (-0.1 to 0.1) | 0.0 (0.0 to 0.1) | 0.0 (-0.1 to 0.0) |
| Month-group interaction: Difference in pre-policy slope in synthetic controls vs BC | 0.0 (-0.2 to 0.1) | 0.0 (-0.1 to 0.1) | -0.1 (-0.2 to 0.0) |
| Level and slope changes after policy implementation in Intervention province and control | | | |
| Immediate change in outcome level at the time of policy change (level change) in BC | -31.2 (-33.0 to -29.3) | 73.2 (71.9 to 74.4) | -41.9 (-43.4 to -40.4) |
| Difference in immediate change in outcome level at time of policy introduction in synthetic controls (vs BC) | 30.6 (28.0 to 33.1) | -73.1 (-74.8 to -71.3) | 41.1 (39.0 to 43.3) |
| Difference in slope between pre-policy and post-policy periods (trend change) in BC | -0.1 (-0.2 to 0.1) | 0.1 (0.0 to 0.2) | 0.0 (-0.1 to 0.1) |
| Difference in trend change in synthetic controls vs BC | 0.1 (-0.1 to 0.2) | -0.1 (-0.2 to 0.0) | 0.1 (0.0 to 0.3) |
| Summary measures: Predicted versus expected counterfactual monthly values in BC and control at 2-years post-policy | | | |
| Monthly difference (predicted minus counterfactual) in BC | -33.2 (-35.8 to -30.7) | 75.3 (73.6 to 76.9) | -41.9 (-45.0 to -38.9) |
| Monthly difference (predicted minus counterfactual) in synthetic control | -1.2 (-3.7 to 1.3) | 0.1 (-1.6 to 1.8) | 3.1 (0.2 to 5.9) |
| Monthly difference of differences (BC - Synthetic control) | -32.0 (-35.6 to -28.4) | 75.2 (72.8 to 77.5) | -45.0 (-49.2 to -40.8) |
| Monthly ratio (predicted divided by counterfactual) in BC | 0.1 (0.1 to 0.2) | 6.4 (5.7 to 7.1) | 0.1 (0.1 to 0.1) |
| Monthly ratio (predicted divided by counterfactual) in Synthetic control | 1.0 (0.9 to 1.0) | 1.0 (0.9 to 1.1) | 1.1 (1.0 to 1.1) |
| Monthly ratio difference (BC RR - Synthetic control RR) | -0.8 (-0.9 to -0.7) | 5.4 (4.7 to 6.1) | -1.0 (-1.0 to -0.9) |
| Other values estimated with bootstrapping | | | |
| Last policy month Synthetic control counterfactual value | 28.0 (25.6 to 30.4) | 14.0 (12.5 to 15.6) | 51.4 (48.7 to 54.0) |
| Last policy month Synthetic control predicted value | 26.8 (25.9 to 27.8) | 14.1 (13.5 to 14.8) | 54.5 (53.4 to 55.5) |
| Last policy month BC counterfactual value | 38.7 (36.3 to 41.1) | 14.0 (12.5 to 15.6) | 46.7 (44.0 to 49.4) |
| Last policy month BC predicted value | 5.4 (4.5 to 6.4) | 89.3 (88.7 to 89.9) | 4.8 (3.6 to 6.0) |

**Supplemental Table 3. Proportions by payer type for contraceptives: Manitoba policy change CITS model estimates and estimated post-policy values**

| Value | Out-of-pocket % (95%CI) | Public payer % (95%CI) | Private insurance % (95%CI) |
|---|---|---|---|
| Pre-policy level and trend: Intervention province and control | | | |
| Outcome level in April 2021 in MB (intercept) | 30.9 (30.5 to 31.3) | 13.9 (13.8 to 14.0) | 50.6 (49.6 to 51.5) |
| Difference in outcome in April 2021 in controls vs MB (group) | -1.2 (-1.8 to -0.6) | -0.3 (-0.4 to -0.1) | 6.0 (4.7 to 7.4) |
| Pre-policy slope in MB | 0.0 (0.0 to 0.1) | 0.0 (0.0 to 0.0) | 0.0 (0.0 to 0.0) |
| Month-group interaction: Difference in pre-policy slope in synthetic controls vs MB | -0.1 (-0.1 to -0.1) | 0.0 (0.0 to 0.0) | -0.1 (-0.1 to 0.0) |
| Level and slope changes after policy implementation in Intervention province and control | | | |
| Immediate change in outcome level at the time of policy change (level change) in MB | -20.7 (-23.3 to -18.1) | 59.2 (57.6 to 60.9) | -34.3 (-38.2 to -30.4) |
| Difference in immediate change in outcome level at time of policy introduction in synthetic controls (vs MB) | 19.8 (16.1 to 23.4) | -59.6 (-61.9 to -57.3) | 35.7 (30.2 to 41.2) |
| Difference in slope between pre-policy and post-policy periods (trend change) in MB | -0.5 (-1.3 to 0.2) | 1.6 (1.0 to 2.1) | -1.9 (-3.0 to -0.8) |
| Difference in trend change in synthetic controls vs MB | 0.7 (-0.4 to 1.7) | -1.4 (-2.2 to -0.6) | 2.0 (0.5 to 3.6) |
| Summary measures: Predicted versus expected counterfactual monthly values in MB and control at last month post-policy | | | |
| Monthly difference (predicted minus counterfactual) in MB | -25.8 (-28.1 to -23.5) | 71.3 (69.5 to 73.1) | -45.5 (-48.7 to -42.3) |
| Monthly difference (predicted minus counterfactual) in synthetic control | -0.8 (-3.0 to 1.5) | -0.3 (-2.1 to 1.5) | 2.3 (-0.8 to 5.5) |
| Monthly difference of differences (MB - Synthetic control) | -25.0 (-28.3 to -21.8) | 71.6 (69.1 to 74.2) | -47.8 (-52.3 to -43.4) |
| Monthly risk ratio (predicted divided by counterfactual) in MB | 0.2 (0.1 to 0.3) | 5.6 (5.3 to 5.9) | 0.1 (0.1 to 0.2) |
| Monthly risk ratio (predicted divided by counterfactual) in Synthetic control | 1.0 (0.9 to 1.1) | 1.0 (0.9 to 1.1) | 1.0 (1.0 to 1.1) |
| Monthly risk ratio difference (MB RR - Synthetic control RR) | -0.8 (-0.9 to -0.7) | 4.6 (4.3 to 4.9) | -0.9 (-1.0 to -0.9) |
| Other values estimated with bootstrapping | | | |
| Last policy month Synthetic control counterfactual value | 27.5 (26.5 to 28.5) | 15.5 (14.7 to 16.3) | 53.0 (51.6 to 54.5) |
| Last policy month Synthetic control predicted value | 26.7 (24.6 to 28.8) | 15.2 (13.6 to 16.8) | 55.4 (52.5 to 58.2) |
| Last policy month MB counterfactual value | 32.6 (31.6 to 33.6) | 15.6 (14.9 to 16.4) | 51.0 (49.6 to 52.3) |
| Last policy month MB predicted value | 6.8 (4.7 to 8.9) | 86.9 (85.3 to 88.6) | 5.5 (2.6 to 8.4) |

**Supplemental Table 4. Costs per capita for contraceptives on BC formulary: CITS model estimates and estimated post-policy values**

| Value | Out-of-pocket<br>$ per capita (95%CI) | Public payer<br>$ per capita (95%CI) | Private insurance<br>$ per capita (95%CI) |
|---|---|---|---|
| Pre-policy level and trend: Intervention province and control | | | |
| Outcome level in April 2021 in BC (intercept) | 0.92 (0.89 to 0.96) | 0.34 (0.27 to 0.41) | 1.31 (1.27 to 1.35) |
| Difference in outcome in April 2021 in controls vs BC (group) | -0.01 (-0.06 to 0.04) | -0.02 (-0.12 to 0.07) | 0.19 (0.14 to 0.24) |
| Pre-policy slope in BC | -0.01 (-0.01 to 0.00) | 0.00 (-0.01 to 0.00) | -0.01 (-0.01 to -0.01) |
| Month-group interaction: Difference in pre-policy slope in synthetic controls vs BC | 0.00 (0.00 to 0.00) | 0.00 (0.00 to 0.01) | 0.00 (0.00 to 0.00) |
| Level and slope changes after policy implementation in Intervention province and control | | | |
| Immediate change in outcome level at the time of policy change (level change) in BC | -0.59 (-0.64 to -0.54) | 1.91 (1.81 to 2.01) | -0.93 (-0.98 to -0.87) |
| Difference in immediate change in outcome level at time of policy introduction in synthetic controls (vs BC) | 0.54 (0.47 to 0.61) | -1.91 (-2.05 to -1.77) | 0.88 (0.80 to 0.95) |
| Difference in slope between pre-policy and post-policy periods (trend change) in BC | 0.00 (0.00 to 0.01) | -0.01 (-0.02 to -0.01) | 0.01 (0.01 to 0.01) |
| Difference in trend change in synthetic controls vs BC | 0.00 (0.00 to 0.01) | 0.02 (0.01 to 0.03) | -0.01 (-0.01 to 0.00) |
| Summary measures: Predicted versus expected counterfactual monthly values in BC and control at last month post-policy | | | |
| Monthly difference (predicted minus counterfactual) in BC | -0.50 (-0.58 to -0.43) | 1.53 (1.38 to 1.68) | -0.72 (-0.85 to -0.58) |
| Monthly difference (predicted minus counterfactual) in synthetic control | 0.10 (0.03 to 0.17) | 0.03 (-0.12 to 0.17) | 0.08 (-0.06 to 0.22) |
| Monthly difference of differences (BC - Synthetic control) | -0.60 (-0.70 to -0.50) | 1.51 (1.29 to 1.72) | -0.79 (-0.99 to -0.60) |
| Monthly risk ratio (predicted divided by counterfactual) in BC | 0.16 (0.11 to 0.21) | 9.37 (-7.67 to 26.40) | 0.13 (0.06 to 0.19) |
| Monthly risk ratio (predicted divided by counterfactual) in Synthetic control | 1.16 (1.03 to 1.29) | 1.27 (-0.41 to 2.95) | 1.09 (0.93 to 1.24) |
| Monthly risk ratio difference (BC RR - Synthetic control RR) | -1.00 (-1.15 to -0.86) | 8.10 (-9.03 to 25.23) | -0.96 (-1.13 to -0.79) |
| Other values estimated with bootstrapping | | | |
| Last policy month Synthetic control counterfactual value | 0.62 (0.55 to 0.68) | 0.24 (0.10 to 0.38) | 0.96 (0.83 to 1.09) |
| Last policy month Synthetic control predicted value | 0.71 (0.69 to 0.74) | 0.27 (0.21 to 0.32) | 1.04 (0.99 to 1.09) |
| Last policy month BC counterfactual value | 0.60 (0.53 to 0.66) | 0.22 (0.08 to 0.36) | 0.82 (0.69 to 0.95) |
| Last policy month BC predicted value | 0.09 (0.07 to 0.12) | 1.75 (1.70 to 1.81) | 0.10 (0.05 to 0.15) |

**Supplemental Table 5. Costs per capita for contraceptives: Manitoba policy change CITS model estimates and estimated post-policy values**

| Value | Out-of-pocket<br>$ per capita (95% CI) | Public payer<br>$ per capita (95% CI) | Private insurance<br>$ per capita (95% CI) |
|---|---|---|---|
| Pre-policy level and trend: Intervention province and control | | | |
| Outcome level in April 2021 in MB (intercept) | 0.91 (0.88 to 0.94) | 0.40 (0.38 to 0.41) | 1.58 (1.55 to 1.61) |
| Difference in outcome in April 2021 in controls vs MB (group) | 0.01 (-0.03 to 0.06) | 0.02 (0.00 to 0.04) | 0.16 (0.12 to 0.21) |
| Pre-policy slope in MB | 0.00 (-0.01 to 0.00) | 0.00 (0.00 to 0.00) | -0.01 (-0.01 to -0.01) |
| Month-group interaction: Difference in pre-policy slope in synthetic controls vs MB | 0.00 (0.00 to 0.00) | 0.00 (0.00 to 0.00) | 0.00 (0.00 to 0.00) |
| Level and slope changes after policy implementation in Intervention province and control | | | |
| Immediate change in outcome level at the time of policy change (level change) in MB | -0.44 (-0.54 to -0.34) | 1.50 (1.44 to 1.57) | -0.70 (-0.86 to -0.54) |
| Difference in immediate change in outcome level at time of policy introduction in synthetic controls (vs MB) | 0.46 (0.32 to 0.60) | -1.51 (-1.60 to -1.43) | 0.75 (0.53 to 0.98) |
| Difference in slope between pre-policy and post-policy periods (trend change) in MB | -0.01 (-0.04 to 0.01) | 0.05 (0.03 to 0.07) | -0.06 (-0.10 to -0.01) |
| Difference in trend change in synthetic controls vs MB | 0.01 (-0.03 to 0.05) | -0.04 (-0.07 to -0.02) | 0.05 (-0.02 to 0.12) |
| Summary measures: Predicted versus expected counterfactual monthly values in MB and control at last month post-policy | | | |
| Monthly difference (predicted minus counterfactual) in MB | -0.52 (-0.59 to -0.46) | 1.83 (1.77 to 1.89) | -0.93 (-1.09 to -0.77) |
| Monthly difference (predicted minus counterfactual) in synthetic control | 0.02 (-0.04 to 0.09) | 0.04 (-0.02 to 0.10) | 0.04 (-0.12 to 0.19) |
| Monthly difference of differences (MB - Synthetic control) | -0.55 (-0.65 to -0.45) | 1.79 (1.71 to 1.87) | -0.97 (-1.19 to -0.74) |
| Monthly risk ratio (predicted divided by counterfactual) in MB | 0.25 (0.16 to 0.34) | 6.23 (5.75 to 6.72) | 0.17 (0.04 to 0.30) |
| Monthly risk ratio (predicted divided by counterfactual) in Synthetic control | 1.04 (0.93 to 1.14) | 1.11 (0.93 to 1.30) | 1.03 (0.90 to 1.16) |
| Monthly risk ratio difference (MB RR - Synthetic control RR) | -0.79 (-0.93 to -0.65) | 5.12 (4.61 to 5.63) | -0.86 (-1.05 to -0.68) |
| Other values estimated with bootstrapping | | | |
| Last policy month Synthetic control counterfactual value | 0.68 (0.65 to 0.71) | 0.33 (0.30 to 0.36) | 1.20 (1.13 to 1.27) |
| Last policy month Synthetic control predicted value | 0.70 (0.64 to 0.77) | 0.37 (0.31 to 0.42) | 1.24 (1.10 to 1.38) |
| Last policy month MB counterfactual value | 0.70 (0.67 to 0.73) | 0.35 (0.32 to 0.38) | 1.12 (1.05 to 1.18) |
| Last policy month MB predicted value | 0.17 (0.11 to 0.23) | 2.18 (2.12 to 2.23) | 0.19 (0.04 to 0.33) |

**Supplemental Table 6. Cash costs per capita in BC by age groups for on-formulary contraceptives: CITS model estimates and estimated post-policy values**

| Value | 15-19y<br>$ per capita (95% CI) | 20-29y<br>$ per capita (95% CI) | 30-39y<br>$ per capita (95% CI) | 40-49y<br>$ per capita (95% CI) |
|---|---|---|---|---|
| Pre-policy level and trend: Intervention province and control | | | | |
| Outcome level in April 2021 in BC (intercept) | 0.91 (0.89 to 0.94) | 1.67 (1.63 to 1.71) | 0.93 (0.90 to 0.96) | 0.45 (0.43 to 0.48) |
| Difference in outcome in April 2021 in controls vs BC (group) | 0.03 (-0.01 to 0.06) | -0.06 (-0.12 to -0.01) | -0.03 (-0.08 to 0.01) | 0.02 (-0.02 to 0.05) |
| Pre-policy slope in BC | -0.01 (-0.01 to -0.01) | -0.02 (-0.02 to -0.01) | -0.01 (-0.01 to 0.00) | 0.00 (0.00 to 0.00) |
| Month-group interaction: Difference in pre-policy slope in synthetic controls vs BC | 0.00 (-0.01 to 0.00) | 0.00 (0.00 to 0.01) | 0.00 (0.00 to 0.01) | 0.00 (0.00 to 0.00) |
| Level and slope changes after policy implementation in Intervention province and control | | | | |
| Immediate change in outcome level at the time of policy change (level change) in BC | -0.59 (-0.63 to -0.55) | -0.96 (-1.02 to -0.89) | -0.64 (-0.69 to -0.60) | -0.39 (-0.43 to -0.35) |
| Difference in immediate change in outcome level at time of policy introduction in synthetic controls (vs BC) | 0.57 (0.52 to 0.63) | 0.89 (0.80 to 0.98) | 0.56 (0.49 to 0.63) | 0.33 (0.27 to 0.38) |
| Difference in slope between pre-policy and post-policy periods (trend change) in BC | 0.01 (0.01 to 0.01) | 0.01 (0.01 to 0.02) | 0.00 (0.00 to 0.01) | 0.00 (0.00 to 0.00) |
| Difference in trend change in synthetic controls vs BC | 0.00 (0.00 to 0.01) | -0.01 (-0.01 to 0.00) | 0.00 (0.00 to 0.00) | 0.01 (0.00 to 0.01) |
| Summary measures: Predicted versus expected counterfactual monthly values in BC and control at last month post-policy | | | | |
| Monthly difference (predicted minus counterfactual) in BC | -0.43 (-0.55 to -0.31) | -0.67 (-0.80 to -0.54) | -0.58 (-0.66 to -0.50) | -0.42 (-0.50 to -0.35) |
| Monthly difference (predicted minus counterfactual) in synthetic control | 0.27 (0.15 to 0.39) | 0.16 (0.03 to 0.28) | 0.02 (-0.06 to 0.11) | 0.08 (0.01 to 0.15) |
| Monthly difference of differences (BC - Synthetic control) | -0.70 (-0.87 to -0.53) | -0.82 (-1.01 to -0.64) | -0.60 (-0.72 to -0.48) | -0.50 (-0.60 to -0.40) |
| Monthly risk ratio (predicted divided by counterfactual) in BC | 0.18 (0.08 to 0.27) | 0.21 (0.14 to 0.28) | 0.14 (0.10 to 0.19) | 0.10 (0.03 to 0.16) |
| Monthly risk ratio (predicted divided by counterfactual) in Synthetic control | 1.70 (1.18 to 2.21) | 1.17 (1.02 to 1.32) | 1.03 (0.92 to 1.15) | 1.20 (0.99 to 1.41) |
| Monthly risk ratio difference (BC RR - Synthetic control RR) | -1.52 (-2.04 to -1.00) | -0.96 (-1.13 to -0.79) | -0.89 (-1.02 to -0.76) | -1.11 (-1.32 to -0.89) |
| Other values estimated with bootstrapping | | | | |
| Last policy month Synthetic control counterfactual value | 0.41 (0.30 to 0.52) | 0.95 (0.84 to 1.07) | 0.73 (0.65 to 0.81) | 0.40 (0.34 to 0.46) |
| Last policy month Synthetic control predicted value | 0.68 (0.63 to 0.72) | 1.11 (1.06 to 1.16) | 0.75 (0.72 to 0.78) | 0.48 (0.45 to 0.50) |
| Last policy month BC counterfactual value | 0.53 (0.41 to 0.64) | 0.85 (0.73 to 0.96) | 0.68 (0.60 to 0.76) | 0.47 (0.40 to 0.53) |
| Last policy month BC predicted value | 0.09 (0.05 to 0.13) | 0.18 (0.12 to 0.23) | 0.10 (0.07 to 0.13) | 0.04 (0.02 to 0.07) |

**Supplemental Table 7. Cash costs per capita by age groups: Manitoba policy change CITS model estimates and estimated post-policy values**

| Value | 15-19y $ per capita (95% CI) | 20-29y $ per capita (95% CI) | 30-39y $ per capita (95% CI) | 40-49y $ per capita (95% CI) |
|---|---|---|---|---|
| Pre-policy level and trend: Intervention province and control | | | | |
| Outcome level in April 2021 in MB (intercept) | 0.83 (0.79 to 0.87) | 1.68 (1.64 to 1.71) | 0.93 (0.89 to 0.97) | 0.50 (0.48 to 0.52) |
| Difference in outcome in April 2021 in controls vs MB (group) | -0.02 (-0.07 to 0.04) | -0.04 (-0.08 to 0.01) | 0.04 (-0.01 to 0.10) | 0.05 (0.02 to 0.08) |
| Pre-policy slope in MB | -0.01 (-0.01 to 0.00) | -0.01 (-0.02 to -0.01) | 0.00 (0.00 to 0.00) | 0.00 (0.00 to 0.00) |
| Month-group interaction: Difference in pre-policy slope in synthetic controls vs MB | 0.00 (0.00 to 0.00) | 0.00 (0.00 to 0.00) | 0.00 (0.00 to 0.00) | 0.00 (0.00 to 0.00) |
| Level and slope changes after policy implementation in Intervention province and control | | | | |
| Immediate change in outcome level at the time of policy change (level change) in MB | -0.39 (-0.56 to -0.23) | -0.70 (-0.83 to -0.56) | -0.50 (-0.62 to -0.39) | -0.41 (-0.50 to -0.33) |
| Difference in immediate change in outcome level at time of policy introduction in synthetic controls (vs MB) | 0.47 (0.24 to 0.71) | 0.75 (0.56 to 0.95) | 0.50 (0.33 to 0.67) | 0.42 (0.30 to 0.54) |
| Difference in slope between pre-policy and post-policy periods (trend change) in MB | -0.01 (-0.05 to 0.04) | 0.01 (-0.03 to 0.04) | -0.02 (-0.05 to 0.02) | -0.01 (-0.03 to 0.02) |
| Difference in trend change in synthetic controls vs MB | -0.01 (-0.08 to 0.05) | -0.01 (-0.07 to 0.04) | 0.03 (-0.01 to 0.08) | 0.03 (-0.01 to 0.06) |
| Summary measures: Predicted versus expected counterfactual monthly values in MB and control at last two years post-policy | | | | |
| Monthly difference (predicted minus counterfactual) in MB | -0.42 (-0.56 to -0.28) | -0.74 (-0.87 to -0.61) | -0.59 (-0.69 to -0.50) | -0.47 (-0.58 to -0.37) |
| Monthly difference (predicted minus counterfactual) in synthetic control | -0.07 (-0.21 to 0.07) | -0.01 (-0.15 to 0.12) | 0.09 (-0.01 to 0.18) | 0.11 (0.05 to 0.17) |
| Monthly difference of differences (MB - Synthetic control) | -0.34 (-0.54 to -0.15) | -0.73 (-0.91 to -0.54) | -0.68 (-0.81 to -0.54) | -0.58 (-0.71 to -0.46) |
| Monthly risk ratio (predicted divided by counterfactual) in MB | 0.28 (0.06 to 0.50) | 0.27 (0.16 to 0.39) | 0.26 (0.15 to 0.37) | 0.15 (-0.03 to 0.33) |
| Monthly risk ratio (predicted divided by counterfactual) in Synthetic control | 0.88 (0.65 to 1.11) | 0.99 (0.86 to 1.11) | 1.12 (0.99 to 1.25) | 1.22 (1.09 to 1.35) |
| Monthly risk ratio difference (MB RR - Synthetic control RR) | -0.60 (-0.91 to -0.28) | -0.71 (-0.88 to -0.54) | -0.86 (-1.03 to -0.69) | -1.07 (-1.29 to -0.84) |
| Other values estimated with bootstrapping | | | | |
| Last policy month Synthetic control counterfactual value | 0.60 (0.54 to 0.66) | 1.06 (1.00 to 1.12) | 0.75 (0.71 to 0.79) | 0.50 (0.48 to 0.53) |
| Last policy month Synthetic control predicted value | 0.53 (0.40 to 0.65) | 1.05 (0.93 to 1.17) | 0.83 (0.75 to 0.92) | 0.61 (0.56 to 0.67) |
| Last policy month MB counterfactual value | 0.58 (0.52 to 0.64) | 1.02 (0.96 to 1.08) | 0.80 (0.76 to 0.84) | 0.56 (0.53 to 0.59) |
| Last policy month MB predicted value | 0.16 (0.04 to 0.29) | 0.28 (0.16 to 0.40) | 0.20 (0.12 to 0.29) | 0.08 (-0.02 to 0.19) |

**Supplemental Table 8. Cumulative total costs by primary payer type in Year 1*, Fiscal year (FY) 2024 and Year 2/FY 2025 based on model-estimated total costs for prescription contraceptives ($ CAD) in BC**

| Actual costs based on IQVIA reported dispensations and cost data (dispensations x unit cost x markup + dispensation fee) | FY 2024 (Year 1) (Apr 2023 - Mar 2024) $CAD Million | Year 1* (May 2023 - Apr 2024) Cumulative cost $CAD Million | FY 2024 (Year 2) (Apr 2024 - Mar 2024) Cumulative cost $CAD Million |
|---|---|---|---|
| BC out-of-pocket measured | 4.45 M | 4.43 M | 3.54 M |
| BC private insurance measured $ | 5.01 M | 5.00 M | 3.80 M |
| BC public payer measured | 34.76 M | 34.08 M | 32.81 M |
| Model-estimated total costs in BC post-policy<br>Total and 95% CI estimated with bootstrapping | | | |
| Out-of-pocket | Not available | 4.34 M 95%CI (4.12 M to 4.57 M) | 3.29 M 95%CI (2.98 M to 3.60 M) |
| Private insurance | Not available | 5.01 M 95%CI (4.67 M to 5.35 M) | 3.74 M 95%CI (3.29 M to 4.20 M) |
| Public insurance | Not available | 34.2 M 95%CI (33.6 M to 34.8 M) | 32.4 M 95%CI (31.8 M to 33.0 M) |
| Model-estimated change in costs relative to counterfactual<br>Total and 95% CI estimated with bootstrapping | | | |
| Out-of-pocket ($) | Not available | -9.58 M 95%CI (-10.2 M to -8.94 M) | -9.75 M 95%CI (-10.7 M to -8.81 M) |
| Private insurance ($) | Not available | -14.65 M 95%CI (-15.6 M to -13.7 M) | -14.6 M 95%CI (-16.0 M to -13.2 M) |
| Public insurance ($) | Not available †estimated: 30.5 M (29 M to 31.9 M) | 30.0 M 95%CI (28.5 M to 31.4 M) | 28.5 M 95%CI (26.4 M to 30.6 M) |

Abbreviations: 95% CI = 95% Confidence interval; CAD = Canadian dollars; FY = Fiscal year; M = Million

* Year 1 model-estimated cumulative costs exclude April 2023 because ITS models excluded this month.
† To estimate total costs for FY 2024 (Apr 2023 to Mar 2024), we added 0.5M to the modeled estimate for Year 1 (May 2023 to Apr 2024) based on visual interpretation of the ITS graphs.

Supplemental Table 9. Synthetic controls model weights for primary models

| Province | Proportion out-of-pocket (cash) | Proportion public insurance | Proportion private insurance | Out-of-pocket costs per capita | Public insurance costs per capita | Private insurance costs per capita | Total costs per capita | SARC costs per capita | LARC costs per capita |
|---|---|---|---|---|---|---|---|---|---|
| Ab | 1.000 | 0.133 | 0.000 | 0.093 | 0.454 | 0.000 | 0.014 | 0.000 | 0.062 |
| Sk | 0.000 | 0.111 | 1.000 | 0.097 | 0.106 | 0.000 | 0.003 | 0.000 | 0.064 |
| On | 0.000 | 0.146 | 0.000 | 0.100 | 0.045 | 0.000 | 0.001 | 0.000 | 0.022 |
| Qc | 0.000 | 0.121 | 0.000 | 0.103 | 0.131 | 0.000 | 0.003 | 0.000 | 0.032 |
| Ns | 0.000 | 0.137 | 0.000 | 0.089 | 0.085 | 1.000 | 0.964 | 1.000 | 0.499 |
| Nb | 0.000 | 0.132 | 0.000 | 0.088 | 0.087 | 0.000 | 0.007 | 0.000 | 0.089 |
| Pe | 0.000 | 0.110 | 0.000 | 0.336 | 0.035 | 0.000 | 0.000 | 0.000 | 0.134 |
| Nl | 0.000 | 0.110 | 0.000 | 0.094 | 0.047 | 0.000 | 0.008 | 0.000 | 0.097 |

Note: in cases where synthetic control weights were based on a single province as the most appropriate control, different control specifications were tested to ensure overall models were robust to control selection.

### SUPPLEMENTAL FIGURES

**Supplemental Figure 1. Proportion of dispensed contraception by payer type using a controlled interrupted time series (CITS) for BC and a synthetic control among only contraceptives on the BC formulary.**

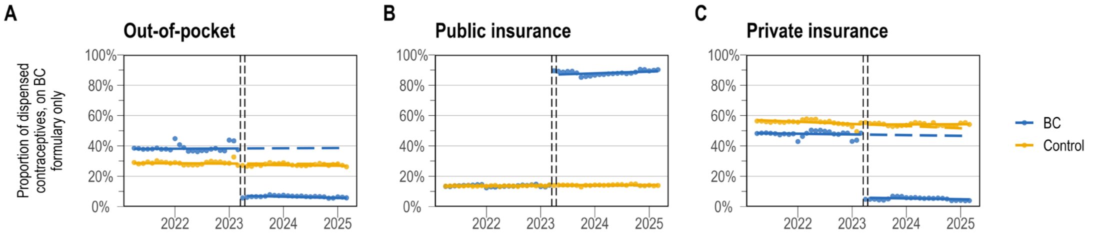


**Supplemental Figure 2. Proportion of dispensed contraception by payer type using a controlled interrupted time series (CITS) for Manitoba and a synthetic control.**

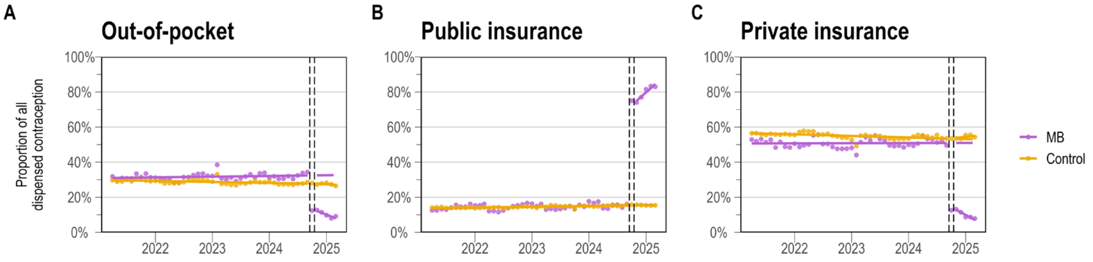


**Supplemental Figure 3. Costs per capita for dispensed contraception by payer type using a controlled interrupted time series (CITS) for BC and a synthetic control among only contraceptives on the BC formulary.**

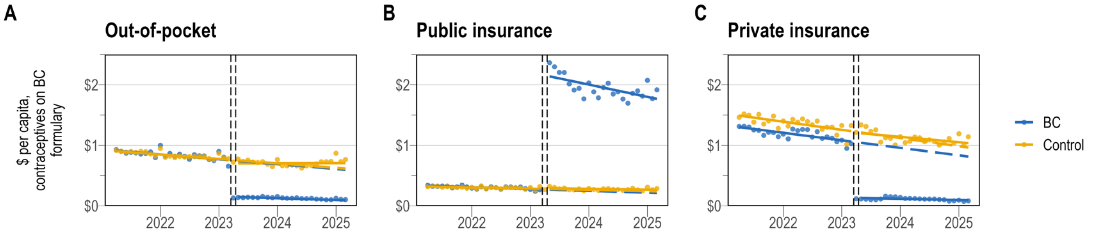


**Supplemental Figure 4. Costs per capita for dispensed contraception by payer type using a controlled interrupted time series (CITS) for Manitoba and a synthetic control.**

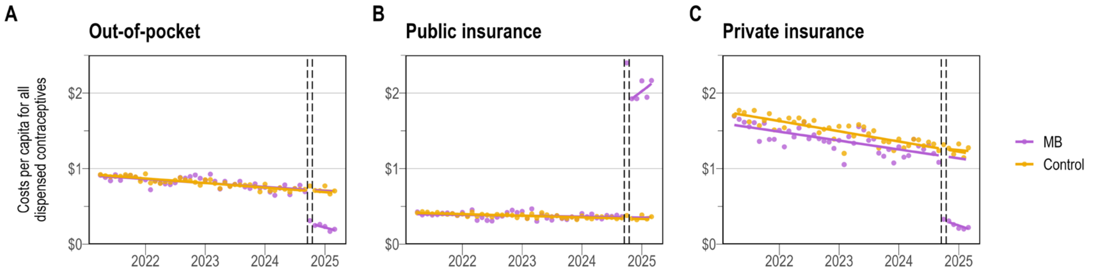

**Supplemental Figure 5. Out-of-pocket costs per capita for on-formulary dispensed contraception by age group using a controlled interrupted time series (CITS) for BC and a synthetic control.**

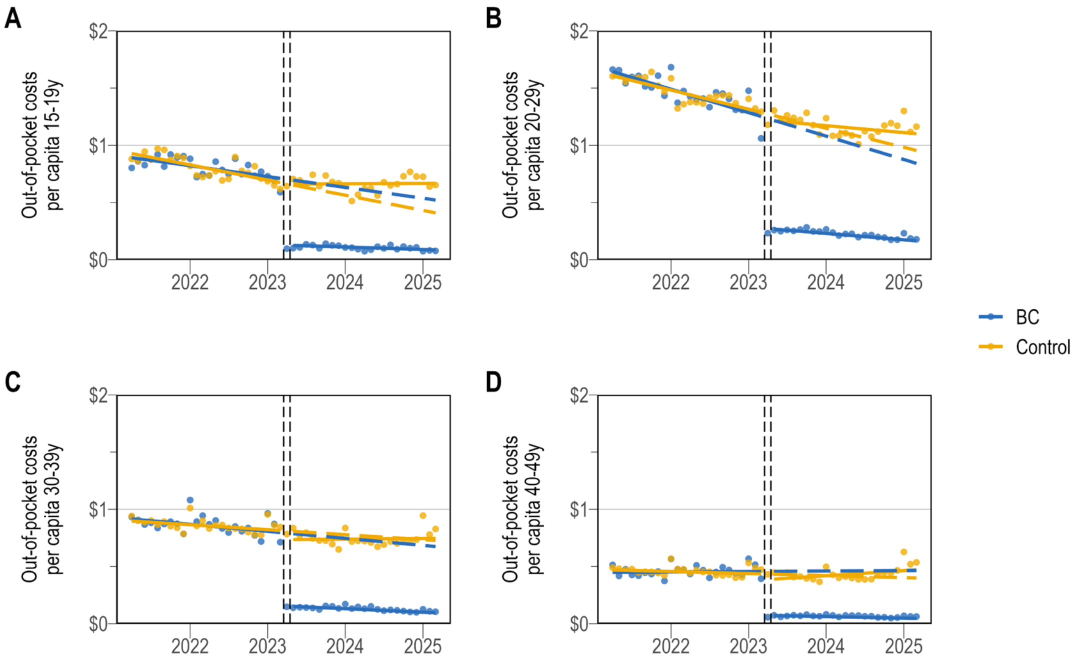


**Supplemental Figure 6. Out-of-pocket costs per capita for dispensed contraception by age group using a controlled interrupted time series (CITS) for Manitoba and a synthetic control.**

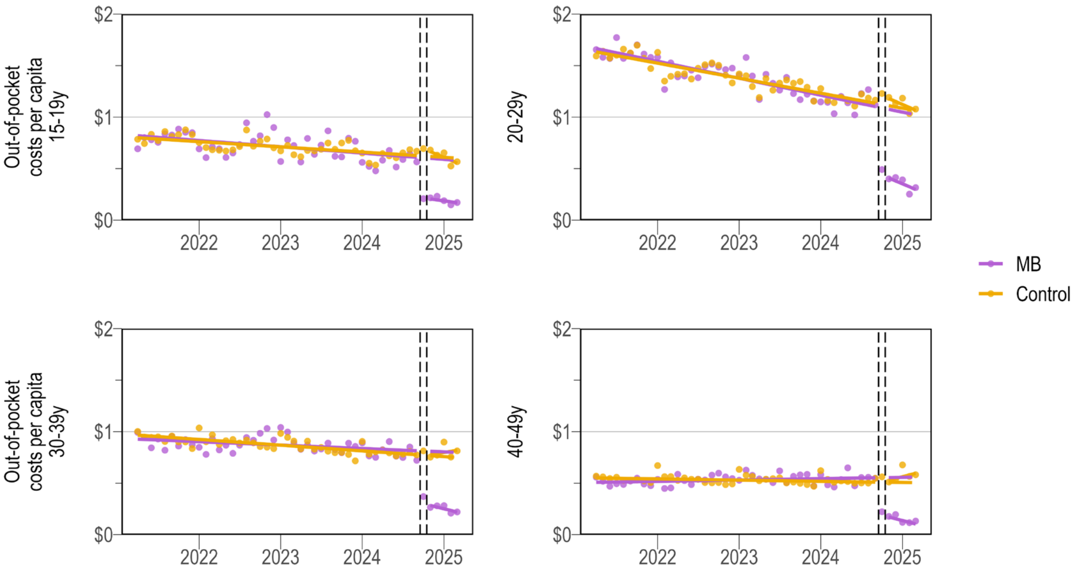

**Supplemental Figure 7. Total costs ($ Million) for prescription contraception costs in all provinces and BC by payer type and ITS model results**

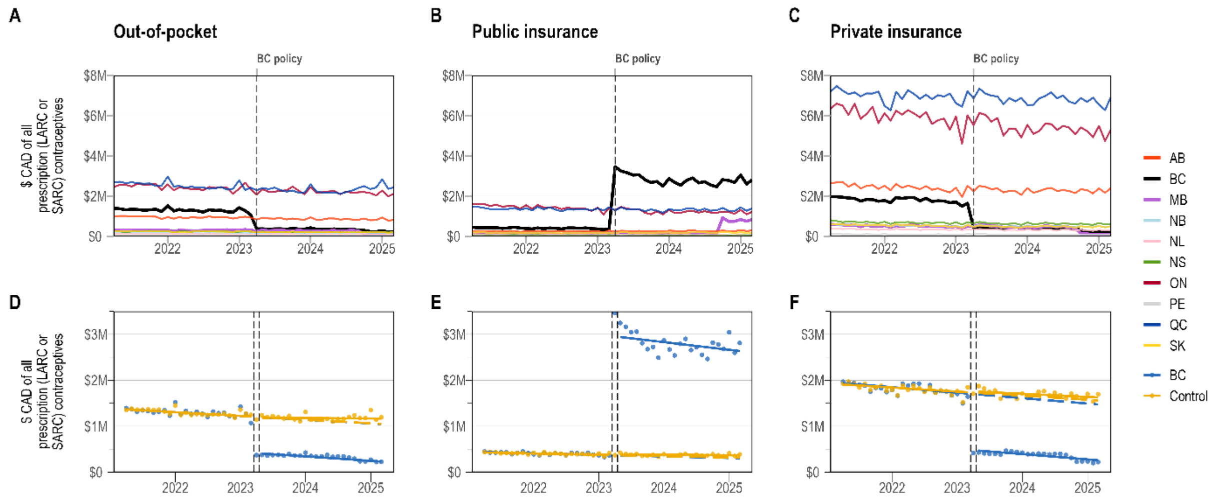
